# Quantifying arbovirus disease and transmission risk at the municipality level in the Dominican Republic: the inception of R_m_

**DOI:** 10.1101/2020.06.30.20143248

**Authors:** Rhys Kingston, Isobel Routledge, Samir Bhatt, Leigh R Bowman

## Abstract

Arboviruses remain a significant cause of morbidity, mortality and economic cost across the global human population. Epidemics of arboviral disease, such as Zika and dengue, also cause significant disruption to health services at local and national levels. This study examined 2014-16 Zika and dengue epidemic data at the sub-national level to characterise transmission across the Dominican Republic.

For each municipality, spatio-temporal mapping was used to characterise disease burden, while data were age and sex standardised to quantify burden distributions among the population. In separate analyses, time-ordered data were combined with the underlying disease migration interval distribution to produce a network of likely transmission chain events, displayed using transmission chain likelihood matrices. Finally, municipal-specific reproduction numbers (R_m_) were established using a Wallinga-Teunis matrix.

Dengue and Zika epidemics peaked during weeks 39-52 of 2015 and weeks 14-27 of 2016 respectively. At the provincial level, dengue attack rates were high in Hermanas Mirabal and San José de Ocoa (58.1 and 49.2 cases per 10,000 population respectively), compared with the Zika burden, which was highest in Independencia and San José de Ocoa (21.2 and 13.4 cases per 10,000 population respectively). Across municipalities, high disease burden was observed in Cotui (622 dengue cases per 10,000 population) and Jimani (32 Zika cases per 10,000 population). Municipal infector-infectee transmission likelihood matrices identified six 0% likelihood transmission events throughout the dengue epidemic and one 0% likelihood transmission event during the Zika epidemic. Municipality reproduction numbers (R_m_) were consistently higher, and persisted for a greater duration during the Zika epidemic (R_m_ = 1.0), than during the dengue epidemic (R_m_ = <1.0).

This research highlights the importance of disease surveillance in land-border municipalities as an early warning for infectious disease transmission. It also demonstrates that a high number of importation events are required to sustain transmission in endemic settings, and vice versa for newly emerged diseases. The inception of a novel epidemiological metric, R_m_, reports transmission risk using standardised spatial units, and can be used to identify high transmission risk municipalities to better focus public health interventions for dengue, Zika, and other infectious diseases.

**Author Summary:** Arboviruses remain a significant cause of morbidity, mortality and economic cost. Between the years 2014-16, two large arbovirus outbreaks occurred in the Dominican Republic. The first was a wave of dengue cases, followed by a large Zika epidemic. Using various mathematical modelling and geospatial approaches, a number of analyses were undertaken to both characterise the pattern of disease transmission and identify high-burden municipalities. Throughout the process, a novel metric was developed: the R_m._ This parameter was used to identify the transmission potential of any given municipality to surrounding municipalities, where >1.0 is high transmission risk, and <1.0 is low transmission risk. This is useful as it provides a standardised approach to determine where public health resources might be focussed to better impact ongoing disease transmission. Additionally, analyses demonstrated the importance of increased disease surveillance in municipalities that share land borders with neighbouring countries, and how relatively few disease importation events can spark and sustain an epidemic.

## Introduction

Arboviruses are an informal name for a group of viruses transmitted by arthropods such as ticks, mosquitoes and sand flies (1) - members of which include Rift Valley Fever, Chikungunya and West Nile Virus (2). Arboviruses are commonly zoonotic, and the cause of increasing human disease worldwide. In recent years, the arboviruses Zika and dengue have afflicted millions via endemic and epidemic transmission, in part due to relatively few, effective means of control (3). Indeed, current estimates suggest that the global annual burden of dengue infections is 390 million, with 96 million manifesting clinically (4). Those at risk number 3.97 billion across 128 countries worldwide (5). In the case of Zika, estimates of the global burden are not yet available, however by the end of 2018, the Pan American Health Organisation (PAHO) had reported 19,020 suspected cases of Zika, with 1,379 laboratory confirmed cases in Brazil alone (6).

Dengue and Zika are principally transmitted via *Aedes* mosquitoes. When a female *Aedes* mosquito bites an infected human, the mosquito ingests a blood meal containing the virus, at which point it enters the midgut, proliferates, and spreads to the salivary glands. Once the mosquito bites another person, the cycle is complete (7). However, vertical transmission can also occur, and while this is relatively rare for dengue (8), such transmission is more common with Zika; indeed in a prospective cohort, 26% of maternal cases resulted in vertical transmission to the unborn foetus (9). Importantly, sexual transmission between humans is also a significant driver of Zika epidemiology (10).

The basic reproduction number (R_0_) describes the average number of secondary infections produced by a single infectious individual in a totally susceptible population (11). Epidemics involving novel pathogens are best described using R_0_, due to the absence of existing population immunity (12). By contrast, R_eff_ is most appropriate in endemic settings (11), when part of the population is already immune (12). In the absence of field data, mathematical modelling is used to average the expected number of new infections over all possible infected individuals. This idea can be represented by a matrix where the reproduction number is recognised as the dominant eigenvalue of an operator, which is linear for every pair of functions, and can be calculated whilst considering other factors such as age stratification (13).

The simplest form of epidemiological modelling is mechanistic, which deploys compartments with interconnected per capita rates to describe the movement of individuals between disease states (14). This field has since been further expanded to include network analysis. Wallinga and Teunis applied this approach (15) to estimate both the serial interval distribution (16) and the R_eff_ of Severe Acute Respiratory Syndrome (SARS). In similar research, Routledge *et al*., 2018 also used network-based analysis to predict malaria elimination time scales (17). Together, these studies further developed mathematical modelling used to calculate individual reproduction numbers (18) while building on the established Reed-Frost model of epidemic transmission (19)(20). And while these approaches are powerful, they are highly reliant on granular data to infer geospatial disease spread at fine scales, yet these data are not always available

Accordingly, this research sought to further analyse data in Bowman *et al*., 2018 (21) by describing the geospatial transmission of dengue and Zika using network-reconstruction and the R_0_ at the regional level.

## Methods

### Country context: Dominican Republic

The Dominican Republic is a country in the Greater Antilles region of the Caribbean (Fig 1).

**Fig 1.**
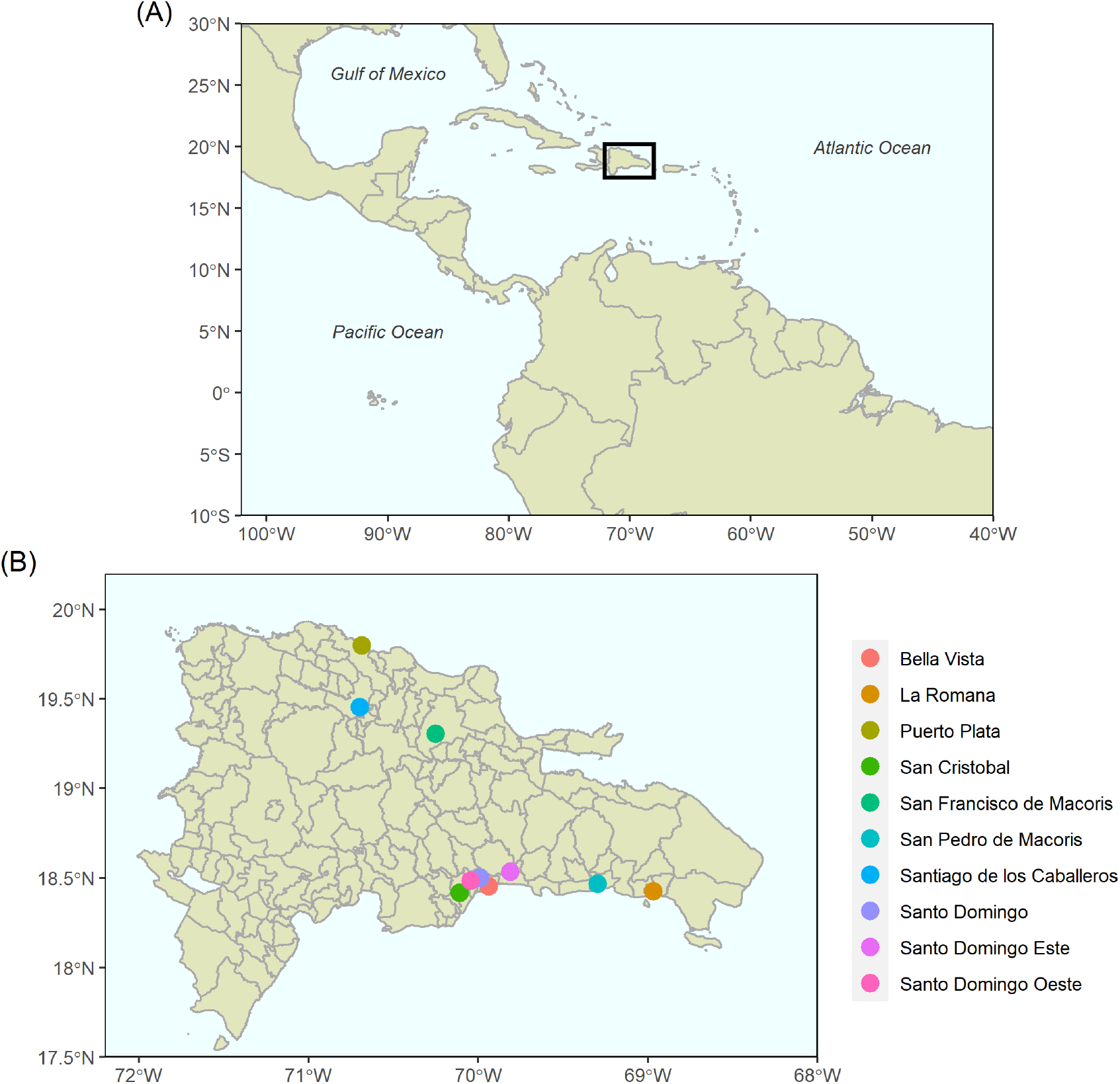
Mapping the Central and South American geographic area. **(A)** The location of the Dominican Republic on a continental scale. **(B)** Administrative municipal boundaries map of the Dominican Republic and ten most populous cities (22). Dominican Republic administrative boundaries, as at 2010, were obtained from the Humanitarian Data Exchange (23). The shapefiles relied on ggplot package to realise the image (24).

It shares an island with Haiti (25), and according to demographic data released by the World Bank in 2016, the total population was 10.63 million over an area of 48.7 km^2^ (26).

The Dominican Republic has a long history of dengue endemicity, and recent research showed that 98% of the Santo Domingo population was seropositive (27). Between 2014-2016, dengue and Zika outbreaks occurred (21), during which large-scale control measures were deployed across the country. A range of clinical and epidemiological data was collected, providing an opportunity to study the passage of the outbreaks at the population level.

### Datasets

Surveillance data capturing cases of dengue, severe dengue and Zika were extracted from the Dominican Republic healthcare database, Sistema Nacional de Vigilencia Epidemiologica, for the years 2014 to 2016. Variables in the dataset included suspected, probable, confirmed cases (28) (29), date of symptom onset, epidemiological week of onset and date of notification. Demographic data were also collected. To capture aggregate dengue disease states, the following outcome variable labels were used: dengue (uncomplicated dengue); severe dengue (complicated dengue); total dengue (complicated and uncomplicated dengue combined). Suspected incident dengue and Zika cases were used to form all outcome variables. Data were de-identified at source, underwent quality control, and cleaned as described in Bowman *et al*, 2016 (30) and Bowman *et al*, 2018 (21). Population census data, stratified by age and sex for the years 2015-2017, were provided by the Oficina Nacional de Estadística. These data were used to standardise attack rate calculations and to categorise data into five-year age bins. All coding and analyses were performed in RStudio version 3.6.0. (31), and all figures produced using the ‘ggplot2’ package (24).

### Mapping

The municipal distribution of cases (per 10,000 population) for each outcome variable was calculated as a function of case counts and population census data (Equation 1). Maps were generated using shape files (23) with administrative boundaries from 2010.

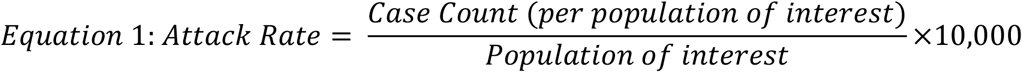

### Statistical analyses

Age and sex standardised attack rates by province were calculated using established methodologies (32). The provincial populations were standardised to the population characteristics of the Dominican Republic using data provided by the Oficina Nacional de Estadística, with 95% confidence intervals calculated using Equation 2:

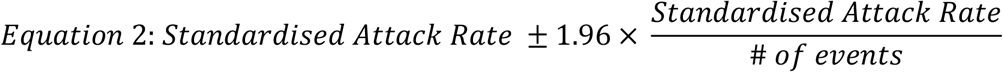

### Disease migration interval distribution

The disease migration interval is a novel parameter, defined in this paper as the time between symptom onset of the infector municipality and symptom onset of the infectee municipality. To calculate the distribution of potential intervals, a matrix of potential migration intervals was calculated by determining non-negative differences between initial symptom onsets within each municipality. The resultant distribution of intervals then informed the probability density of infector, *i*, transmitting infection to the infectee, *j*. This interval reflects a higher order version of the serial interval, which specifies the interval between symptom onset of the infector and infectee individual pairs. The benefit of using the serial interval in estimates is the ability to account for other important distributions of time in the transmission cycle, including the time from symptom onset to infectiousness, intrinsic incubation period, extrinsic incubation period and mosquito transmission rate (17). The probability density of the disease migration interval was fitted to an exponential distribution after visualisation of the data strongly indicated an exponential trend. Fitting the distribution was achieved using maximum likelihood estimation with the exponential maximum likelihood estimator, seen below, where 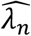 is the maximum likelihood estimator, *n* is the number of independent observations, *x* is a variable from an independent and identically distributed sample and 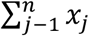 is the sum of all observations. The resultant simulated distribution was used to calculate the Wallinga-Teunis matrix (Equation 3)

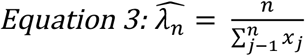

### Determining the transmission likelihood and network

The cases with the earliest symptom onset of dengue or Zika recorded within each municipality were identified, resulting in one case representing the earliest infection event for each municipality. These were ordered by date of symptom onset for each municipality, with no indication of the transmission chain present. Combination of this time-ordered data with a simulation of the underlying disease migration interval distribution produced a network of the most likely transmission chain events. This was achieved by analysing a network of all potential pairwise infector-infectee municipality pairs, and their transmission likelihoods, to isolate the most likely chain of transmission events across municipalities. The network of potential infector-infectee municipal pairs and their transmission likelihoods make up the Wallinga-Teunis matrix (15), made with the ‘IDSpatialStats’ package (33). The matrix itself represents likelihood-based estimation of who-infected-whom using observed dates of initial symptom onset of each municipality. Each square provides the relative likelihood, *p*_*ij*_, that the infector municipality, *i*, has infected an infectee municipality, *j*, given the time difference in symptom onsets of each municipality, *t*_*i*_ *- t*_*j*_. This time difference is captured by the disease migration interval distribution. As such, the relative likelihood that an infectee municipality, *i*, has been infected by an infector municipality, *j*, is the likelihood of this pair, normalised by the likelihood that the infector municipality, *i*, is infected by any other municipality, *k*. This analysis is based around the theory that infection events between the potential pairs follow an independent cascade model (34), where the upper triangular likelihood of the matrix represents all the realistic pairwise transmission likelihoods of the infector-infectee municipal pairs (15).

### Estimating time-varying municipal-specific reproduction numbers

Municipal-specific reproduction numbers (R_m_) were established using the produced Wallinga-Teunis matrix wherein each column represents an infector municipality and each row represents an infectee municipality. To calculate the R_m_ for an infector municipality, *j*, we sum over all infectee municipalities, *i*, weighted by the relative likelihood that the infectee municipality, *i*, has been infected by the infector municipality, *j*. At a municipal level, this reflects the number of municipalities that the infector municipality will go on to infect. The time-varying R_m_ was plotted over time and a Generalised Additive Model smoothing spline was fitted to the data to determine trends and smooth data noise.

### Ethics

Ethical clearance was granted by the Pan American Health Organization Ethics Review Committee (PAHO-ERC; Ref No. 2014-10-0023) and accepted by Dominican Republic Ministry of Health. De-identified and aggregated data were used throughout the study, no further ethical clearance was required.

## Results

### Geostatistical Mapping

Spatial mapping of Zika and dengue was used to determine the highest burden areas for all disease outcomes across the Dominican Republic (Fig 2). Spatio-temporal mapping displays incidence per 10,000 population (non-standardised), while attack rates were standardised according to age and sex with accompanying confidence intervals. These can be seen in S1 Table, for total dengue, and S2 Table, for Zika.

**Fig 2.**
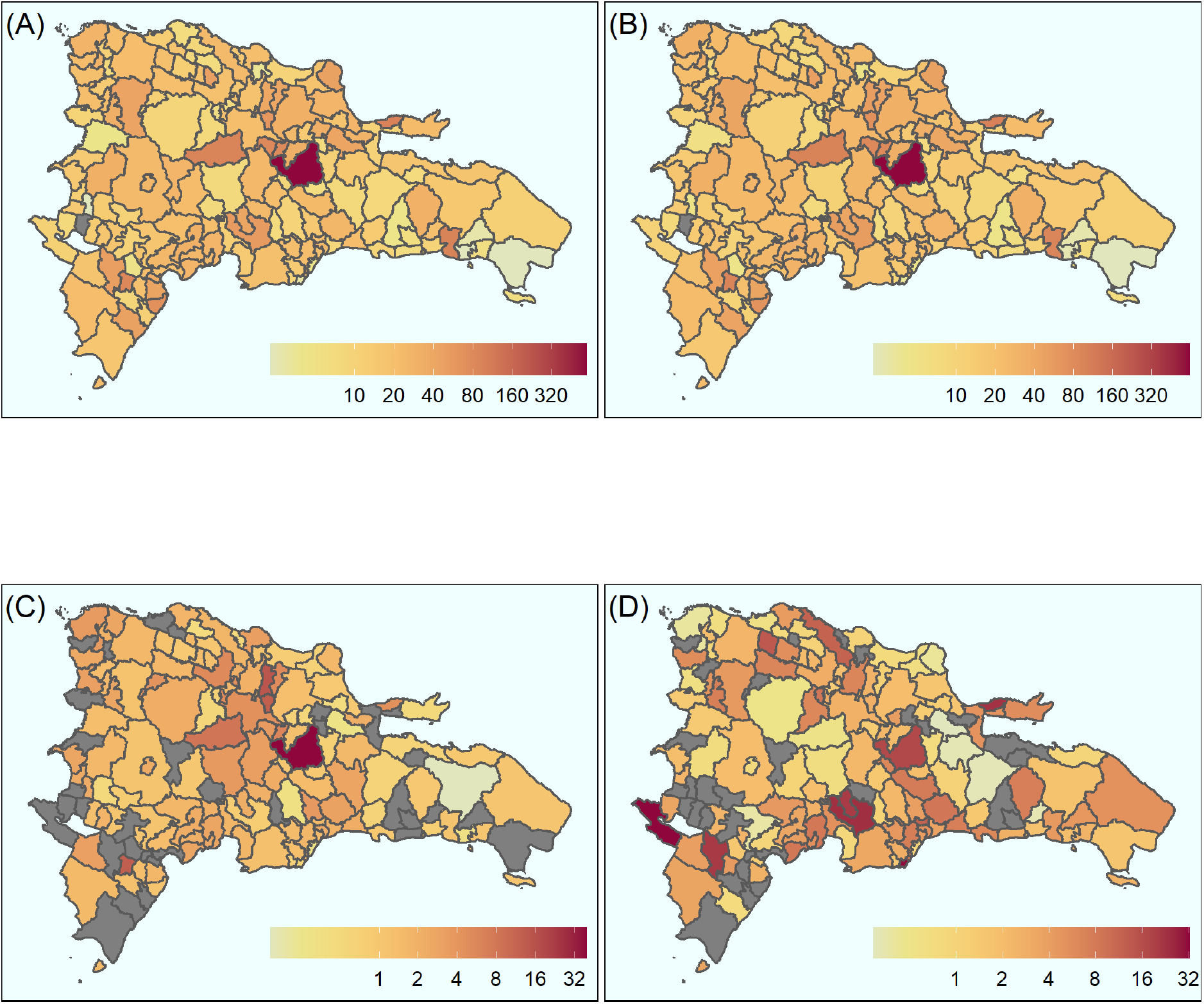
Aggregated spatial distribution of cases over 2014-2016 epidemic period at the municipal level. **(A)** Uncomplicated dengue, **(B)** Dengue, **(C)** Complicated dengue and **(D)** Zika. Continuous colour scale from white (lowest) through to red (highest) for all images, scales vary. Grey areas indicate no data for those municipalities. All counts per 10,000 population.

The municipality of Cotuí recorded the highest burden for each dengue outcome: 583 (uncomplicated dengue), 39 (complicated dengue) and 622 (dengue) cases per 10,000 population respectively (Fig 2), which equates to the largest dengue burden of any municipality. Municipalities that also recorded high dengue burden include Las Terrenas, Jarabacoa and Las Salinas recorded 99, 98 and 95 per 10,000 population respectively (Fig 2B). The burden of complicated dengue was also high in Salcedo, Villa Tapia and Jarabacoa with 15, 12 and 8 cases per 10,000 population respectively (Fig 2C). The highest burden of Zika incident cases was recorded in the west of the country, Jimaní, with 32 cases per 10,000 population (Fig 2D).

All four outcomes were also displayed over time and space, as seen in Figure 3. Where the outcome was dengue, the highest burden municipalities were Jarabacoa, Ramón Santana and La Cienega with 26, 23 and 20 cases per 10,000 population respectively (Fig 3A), which occurred between epidemiological weeks 39-52 in 2015. The highest burden of complicated dengue incident cases was recorded in Las Salinas, Villa Tapia and Jarabacoa with 2 cases per 10,000 population each (Fig 3B) within the same time period.

**Figure 3.**
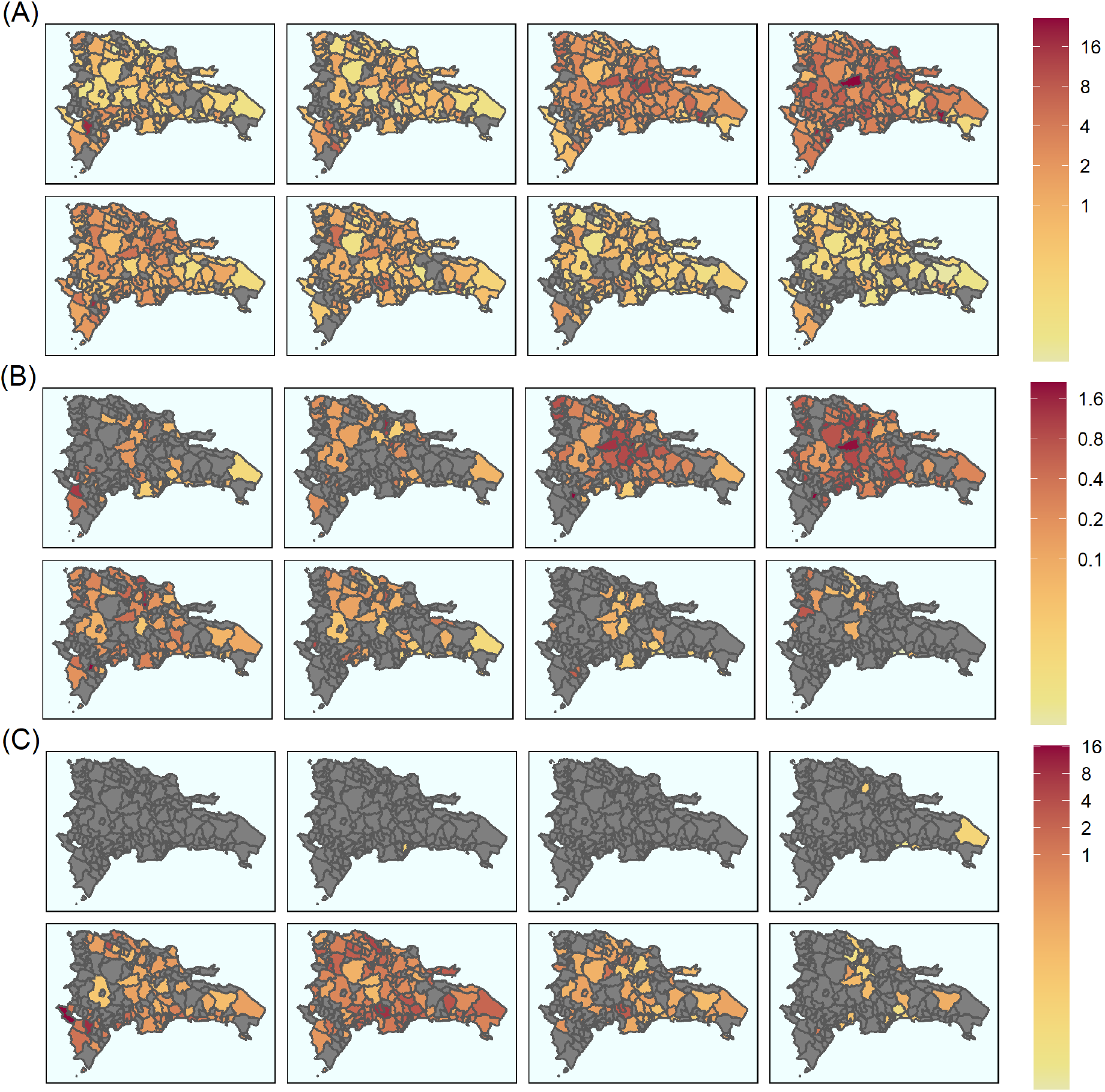
Breakdown of aggregated spatial distribution of cases over 2014-2016 epidemic period by epidemiological week at the municipal level. **(A)** Dengue cases over 2015 (top row) and 2016 (bottom row) **(B)** Complicated dengue cases over 2015 (top row) and 2016 (bottom row). **(C)** Zika cases over 2015 (top row) and 2016 (bottom row). Continuous colour scale from white (lowest) through to red (highest) for all images. Scales vary as shown. Grey areas indicate absence of data. All counts per 10,000 population. Dates are over epidemiological weeks 1-52 for each year, where year is split into weeks 1 – 14, 14 – 27, 27 – 39, and 39 - 52.

The first suspected cases of Zika in 2015 were identified during epidemiological weeks 14 – 27 and reported in San Cristobal (Fig 3C), while the highest burden of Zika disease was reported in Jimaní (16 cases per 10,000 population: epidemiological weeks 1-15). In 2016, the peak of Zika cases occurred between epidemiological weeks 1 – 27 (Fig 3C). Throughout weeks 14 -27, the greatest burden of Zika disease was recorded in Sabana Grande de Palenque, San José de Ocoa and Sabana Larga (13, 9 and 6 cases per 10,000 population respectively) (Fig 3C).

### Transmission Dynamics

A distribution of the disease migration interval, defined as the serial interval calculated between municipalities, instead of individuals, was calculated to capture the spatio-temporal disease dynamics of the dengue and Zika epidemics.

Each municipality was described as either infector or infectee, and the disease migration interval referenced the time between symptom onsets in each of the infector-infectee pairs. The results indicated an exponential distribution between the probability density of secondary municipality symptom onset (infectee) and the time of symptom onset in the infector municipality (Fig 4) for both dengue and Zika. Probability of transmission from infector to infectee municipalities was elevated (∼0.1) for both dengue and Zika at the beginning of the epidemic. Probabilities for dengue remained elevated for the first ∼50 days before tailing off, whereas probabilities for Zika were high for the first ∼125 days before a gradual decline.

**Fig 4.**
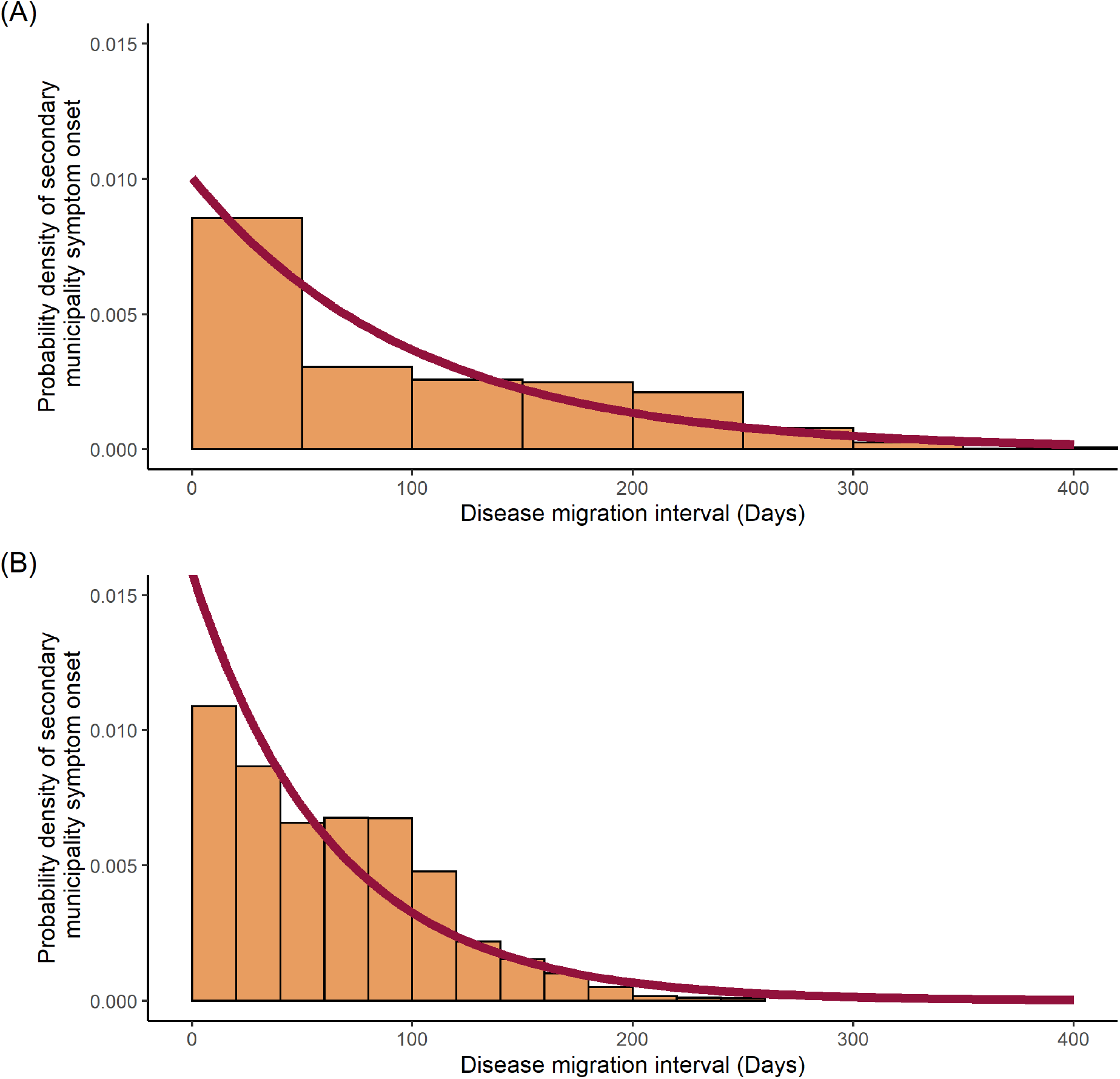
Disease migration interval distribution fitted to exponential distribution. **(A)** Dengue cases, **(B)** Zika cases. Probability density histograms plotted of the disease migration interval represent the distribution of time differences between initial symptom onset within each municipality. Red line represents the maximum likelihood estimation of the exponential distribution describing the data.

The mean disease migration interval, probability of transmission per day (expressed as a rate), standard deviations and log likelihoods for the fitted distributions can be seen in Table 1. The rate for the dengue distribution fitted to 0.01004 (s = 0.00009095) and the rate for Zika distribution was 0.01580 (s = 0.0001793).

**Table 1:** Fitted disease migration interval distribution parameters. Mean disease migration interval calculated by the inverse of the estimated rate

| Parameter | Disease |  |
| --- | --- | --- |
|  | Dengue | Zika |
| Mean Disease Migration Interval (days) | 99.55 | 63.28 |
| Rate (probability of transmission/day) | 0.01004 | 0.01580 |
| Standard Deviation | 0.00009095 | 0.0001793 |
| Log Likelihood | -66950 | -39670 |

The disease migration intervals were used to produce the Wallinga-Teunis matrices (Fig 5) along with the transmission network between infector-infectee municipal pairs for dengue and Zika (Fig 6).

**Fig 5.**
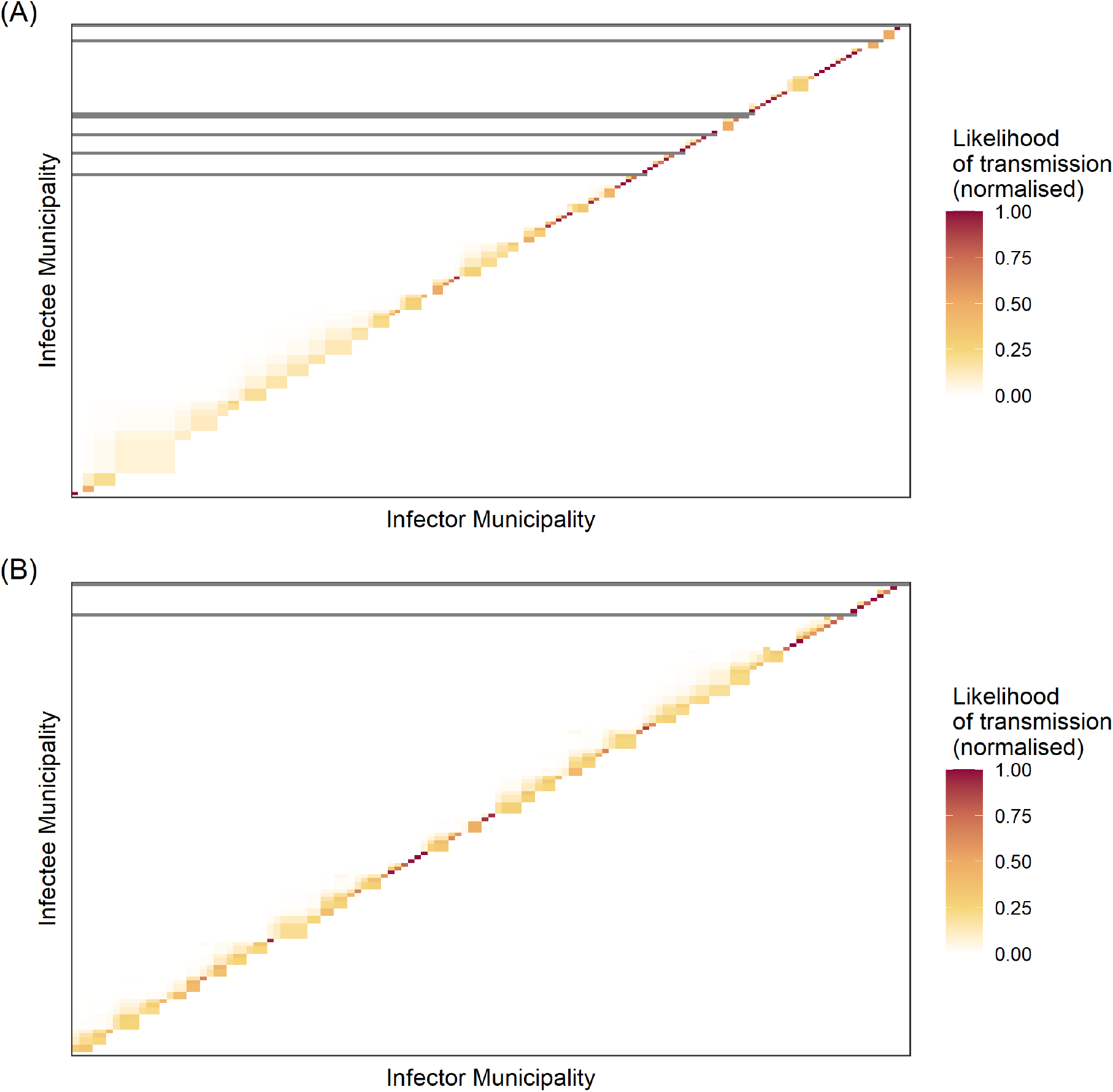
Heatmaps of transmission likelihood of infector-infectee municipal pairs. **(A)** Dengue cases, **(B)** Zika cases. Heatmap *X* axis represents all possible infector municipalities ordered by time of initial symptom onset date; *Y* axis represents all possible infectee municipalities ordered by time of symptom onset. Each square represents the transmission likelihood for said infector-infectee pair. Continuous scale from grey (0) to red (1) represents the normalised likelihood of transmission, with grey squares indicating no likelihood of transmission. Dark grey lines represent where infectee municipalities were unlikely to be infected by other observed municipalities, and so infection occurred by unobserved disease migration.

**Fig 6.**
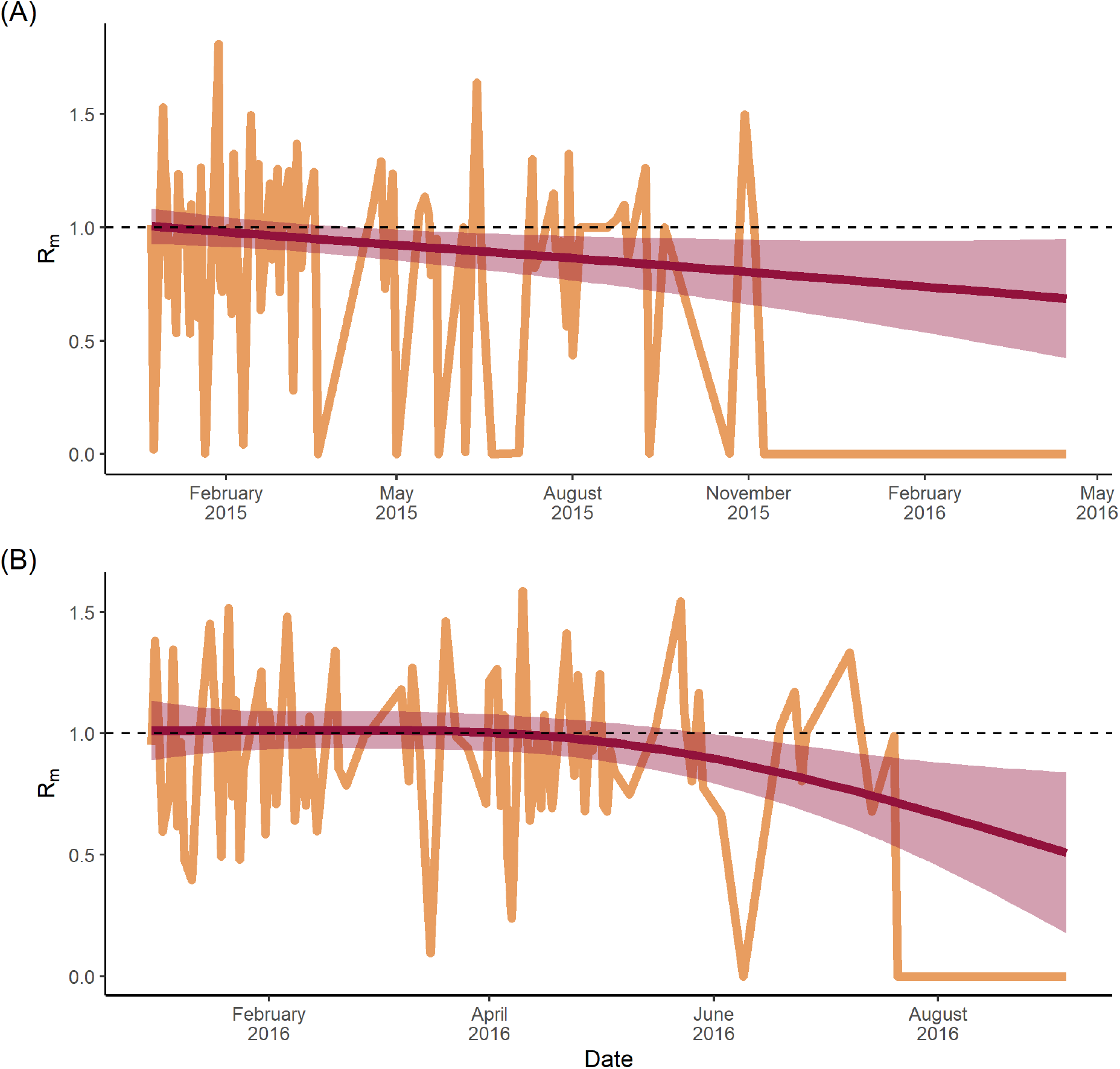
Municipal-specific, time-varying reproduction numbers. **(A)** Dengue cases, **(B)** Zika cases. R_m_ is the number of municipalities a given infector municipality is likely to infect. Red line represents the fitted Generalised Additive Model with smoothing splines and the 95% confidence interval seen as a shaded grey area. Dashed line represents an R_m_ = 1.

During the exponential phase of the outbreak, the transmission likelihood matrix for dengue was populated by lower likelihood transmission events compared with the downward curve (Fig 5A). The matrix also identified six transmission events with a 0% likelihood of transmission between infector and infectee, all of which occurred in the latter half of the transmission network. By comparison, the Zika transmission likelihood matrix (Fig 5B) displayed one transmission event with a 0% likelihood of transmission between infector and infectee, both of which also occurred at the end of the transmission network. The transmission chain itself, however, was populated with many high likelihood transmission events, with few pairwise infector-infectee likelihoods below 50%. In other words, the likelihood of transmission between infector and infectee municipalities increased over time for both dengue and Zika, although probabilities were higher, earlier for Zika.

To obtain the time-varying R_m_, the sum of the transmission likelihoods for each infectee municipality was calculated and plotted over time (Fig 6). For dengue (Fig 6A), there was a linear trend starting at an R_m_ of ∼1.0 in January 2015, which decreased to a value of approximately 0.7 by April 2016. Zika (Fig 6B) showed a consistent R_m_ of approximately 1 until just after April 2016 when it began to decline. By September 2016, the R_m_ was ∼0.4. Grey areas in Figure 6 represent 95% confidence intervals, which increase in size over time and correlate with greater uncertainty as caseload declines.

## Discussion

This research set out to explore the spatio-temporal trends of both dengue and Zika epidemics between 2014-2016, and better define disease progression at a municipality level across the Dominican Republic. Retrospective analysis of incident case data was used to map the spatio-temporal distribution of cases. Transmission likelihood matrices for infector-infectee pairs were generated, and the temporal trend of R_m,_ was calculated to better understand transmission dynamics over time.

Dengue and Zika attack rates over the entire epidemic period varied substantially across the country, likely a result of known transmission drivers (35). As shown in Fig 3A, the peak of the dengue epidemic occurred during epidemiological weeks 39-52 of 2015, which coincided with the implementation of control efforts, such as fogging and public health campaigns (36), that may have stymied transmission (37). By contrast, the peak of the Zika epidemic occurred between weeks 14-27 of 2016 (Fig 3D). Uncomplicated dengue attack rates were highest in the municipality of Cotuí, at 622 cases per 10,000 population. No other municipality recorded >100 cases per 10,000 population. By contrast, the highest Zika attack rate was recorded in the municipality of Jimaní, at 32 per 10,000 population, and equates to a ∼20-fold difference in incidence, demonstrating the continuing burden of dengue in the Dominican Republic.

That Jimani recorded the highest Zika burden in the country is important not only due to the relatively high caseload. Jimani has a population of 400,000 and shares a land border with Haiti. It has undergone rapid expansion in recent decades and is a hub for the movement of people and goods across the border (38). Considering the detection of Zika in Haiti as early as 2014 (39), and that Jimani has become a gateway for larger campaigns in Haiti (40), it is plausible that a number of Zika importation and re-importation events occurred across both sides of the border. This narrative is hypothetically confirmed by spatio-temporal mapping of Zika, showing that western-most municipalities were affected greatly in the early phases of the epidemic, while central and eastern regions were affected later. In light of this, increased disease surveillance capacity in Jimaní could offer valuable early warning for disease events across both sides of the border.

### Human Mobility and Infrastructure

Human movement between neighbourhoods and commuter cities is known to intensify dengue transmission (41) (42). Indeed, those provinces (Hermanas Mirabal, Sánchez Ramirez and La Vega) that share these characteristics reported relatively high uncomplicated dengue attack rates of 58, 48 and 38 cases per 10,000 population respectively (standardised for age and sex) (Table S1). For Hermanas Mirabal and La Vega, high standardised attack rates correlate with their geographical location. They are connected by primary roads DR-132 and DR-1 to San Francisco de Macoris and Santiago De Los Caballeros respectively, two of the ten largest cities by population (22)(43). Sánchez Ramirez province, which includes Cotuí municipality, also places on one of the Dominican Republic’s primary roads (DR-17) between Santo Domingo and San Francisco de Macoris (43). Accordingly, municipalities and towns along major commuter belts would likely benefit from greater surveillance and public health capacity.

### Transmission Chains

The disease migration interval, seen in Fig 4, describes the likelihood of secondary (municipality) infection as a function of the distribution of disease migration intervals for dengue and Zika. This reflects both the infectious period and human mobility. For dengue, the migration interval was heavily skewed towards the first 50 days after symptom onset, in contrast to Zika, which showed a broader distribution over the first 125 days. Given that both pathogens are transmitted via the same *Aedes* vectors, this suggests a more significant role for sexually transmitted Zika (44), at least in terms of transmission drivers throughout the first half of the epidemic.

Wallinga and Teunis (15) first proposed transmission likelihood matrices to identify breaks in transmission chains. In real terms, this equates to the importation of cases, better known as importation events (17). The international and intra-national movement of people, and the influence of asymptomatic or unreported individuals, can be captured using this methodology, which can help identify both the index case and the source of importations/ reintroductions (17). Using absolute dengue cases, this study identified six events that had 0% likelihood of transmission between infector and infectee municipalities, in other words, six importation events (Fig 5A). These occurred during the latter half of the outbreak and constituted a greater number of importation events than the single event observed for Zika. While it is not possible to tease out the origin of each event, the relative frequency of importation events between diseases is not unexpected, given the assumed largely Zika seronegative international population (due to the novel nature of the virus) vs. the global endemicity of dengue (4). However, it is also possible that these events are related to intra-national introductions, where individuals become infected through inter-municipal contacts, reinforcing the importance of human mobility as a transmission driver, but also potentially through asymptomatic or unreported infections. The implicit assumption here is that a lower R_m_ requires a greater number of importation events to sustain transmission, and vice versa.

The likelihood of transmission increased with time from symptom onset for both Zika and dengue. Heatmaps for dengue (Fig 5A) showed increasing likelihood of transmission between infector-infectee pairs, as a function of symptom onset over time, likely indicating multiple smaller importation events in pockets of less well-connected municipalities in rural areas. By contrast, Zika heatmaps demonstrated a more consistent chain of transmission, most likely reflecting a continuous supply of susceptibles infected by two modes of transmission. This is not atypical for Zika, and has been observed in Rio de Janeiro, Brazil, where multiple introductions over a short space of time led to a national crisis (45), since corroborated by phylogenetic analysis linking the strain to French Polynesia (46).

### Municipal Reproduction Number (R_m_)

Defining transmission chains and generating time-varying reproduction numbers can provide epidemiologists with valuable information that inform surveillance, control and response. Methodologies used to generate these metrics are established (47) (48) and have been used to determine the impact of cattle culls on foot-and-mouth disease in the UK (47). However, only a small proportion of such probabilistic studies have focussed on arboviruses, with Salje *et al*., 2016 looking specifically at the transmission dynamics of chikungunya (49). Independent cascade models (34) have also been used to determine interactions across networks for infectious disease outbreaks, yet these focussed on individuals’ data (17), or were used in the context of social network modelling (50). Where this study expands the field is in the use of widely available data at a standard geospatial unit – the municipality – to understand transmission dynamics of infectious diseases, using a newly-defined variant of the basic reproduction number: R_m_. The metric reflects the average number of secondary infectee municipalities arising from a primary infector municipality, and can be interpreted as follows: a municipality with an R_m_ <1 reflects a lower likelihood of infection to other municipalities, while on the other hand, an R_m_ >1 represents increased likelihood of infection to other municipalities.

In this study, dengue R_m_ was recorded as ∼1.0 at the start of the epidemic, but immediately and steadily declined throughout (Fig 7A), likely reflecting two factors: 1) that there was a relatively small pool of susceptibles among a highly mobile population in the early phases of the outbreak and 2) that this pool depleted fairly rapidly as transmission spread from major urban areas before fading throughout less mobile populations. By contrast, the Zika R_m_ (Fig 7B) remained constant at a value of 1.0 for four months between January and April, indicating a large pool of susceptibles (51) that were infected steadily as the infection spread throughout the population. Then in May, the R_m_ steadily declined below 1.0, suggesting both a declining pool of susceptibles and a lower force of infection, perhaps as the virus reached poorly connected rural areas. This transmission pattern has been observed previously in French Polynesia and the Federated States of Micronesia, where high seroprevalence of IgM antibodies in the local population suggested an acute outbreak that infected three quarters of the population over a similar time scale: four months from April – July 2007 (52), before tailing off.

The dengue R_m_ observed in this study provides evidence that vector-borne disease spread between administrative locations, and so between populations, can still occur when the effective municipal reproduction number is below 1. Conversely, Zika, as a newly emerged infection, never recorded an R_m_ of 1.5 or higher, but maintained a consistent R_m_ of approximately 1.0 during the growth and exponential phases of the epidemic. This indicates that even in the presence of a huge pool of mobile susceptibles, vector-borne and sexually transmitted diseases have an R_m_ ‘celling’, pre-determined by the mode of transmission.

This novel metric clearly has benefit. It operates at a scale that broadly aligns with existing geospatial data collection, thus addressing fundamental issues surrounding data and spatial heterogeneity described elsewhere (30, 53, 54). Operationally, the R_m_ can be used to identify high-burden and high-risk municipalities that necessitate intervention, thereby aligning with early warning and response systems that operate on similar spatial scales (30, 55). But it should be cautioned that the R_m_ should be used as a floating metric to guide intervention, rather than a binary threshold used to trigger intervention.

## Limitations

Estimation of the time-varying R_m_ across the Dominican Republic required the heuristic determination of an optimal distribution to describe the disease migration interval distribution. This used the maximum likelihood estimation of an exponential distribution fit to the data, as seen in Figure 4, which required the assumptions that the data was identical, independent, and discrete, and fitted the interval probability density distribution well. This was supported by the log likelihood for each of these models, which were significantly negative, as shown in Table 1, implying a good fit to the data. However, this and the standard deviation for Zika were lower than in dengue, implying the model fitted the dengue epidemic data better than for Zika. This could have been a product of more uncertainty in the exact distribution for Zika due to the smaller sample size of disease migration intervals.

As suspected cases were used in all analyses, there was the potential for misclassification between not only dengue and Zika, but also within the clinical spectrum of dengue, as well as misreporting. All rates were calculated using 2016 census data, so there will be small discrepancies in precision when standardising earlier datasets. Data paucity was an issue for those over 80 years of age resulting in increased noise and less reliability across results within this age group.

Wallinga-Teunis matrices rely on the temporal product of the disease migration interval distribution, so for these analyses, the distribution was reasonably assumed to be exponential. Furthermore, the matrices themselves are dependent on the completeness of the dataset regarding asymptomatic and unreported infections. Consequently, the clarity of the transmission chain could be honed by incorporating predictions on rates of asymptomatic or unreported cases.

The production of time-varying reproduction numbers assumed complete susceptibility to the viruses within the population, which was valid for Zika, but less so for dengue, although susceptibility to the serotype across the population was patently high.

## Conclusions

The results of this study characterised the Zika and dengue burden at the municipality level in the Dominican Republic across 2014-16. Concentrated disease burden within specific municipalities is likely due to the presence of significant transport arteries, both within Dominican Republic and across the border to Haiti, as a conduit for increased human movement and disease dispersal. Therefore, increased surveillance of both vector and epidemiological data (51), alongside targeted public health measures in these municipalities, is needed.

Furthermore, this research highlights the inception of a novel metric used to quantify and determine transmission chains at the municipal level, which can be used to characterise municipality risk, in terms of secondary transmission to neighbouring municipalities. This approach can be generalised to countries worldwide, for multiple infectious diseases, towards refining public health responses by targeting municipalities that are significant contributors to disease spread.

Finally, this study further reinforces the importance of importation events that drive transmission where R_m_ is below one, and conversely the significance of immune-naïve populations in facilitating disease spread, which require fewer importation events to sustain transmission. Future research should be focused on the refinement of these novel metrics, and application of these to characterise municipalities based on a risk-system, reflecting variation in R_m_ outputs.

## Data Availability

All raw data files are available from the Open Science Data Framework: DOI 10.17605/OSF.IO/VYN2B.

https://osf.io/VYN2B/

## Acknowledgments

The authors would like to acknowledge Dr Ronald Skewes-Ramm, Prof Axel Kroeger and Prof Piero Olliaro for their work towards obtaining the original dataset.

## Supplementary Material

**Table S1.**
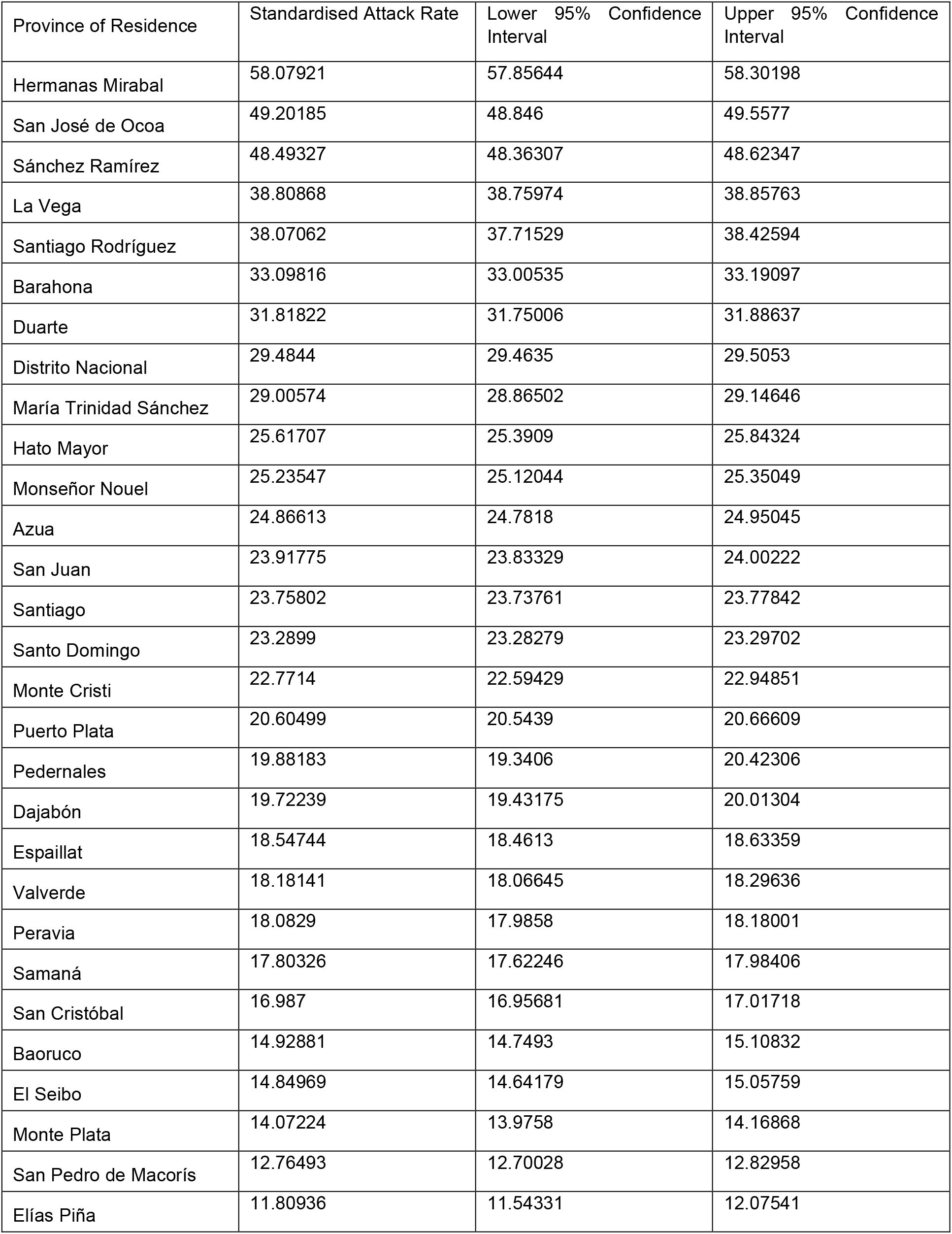
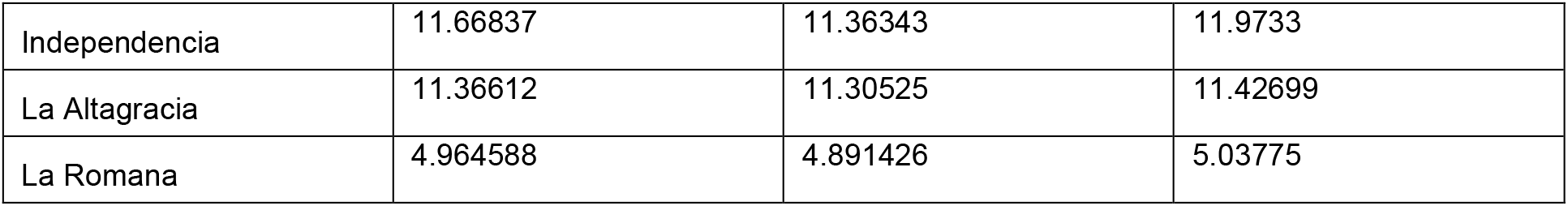
Age and sex standardised attack rates of dengue by province. Respective lower and upper 95% confidence intervals are shown.

**Table S2.**
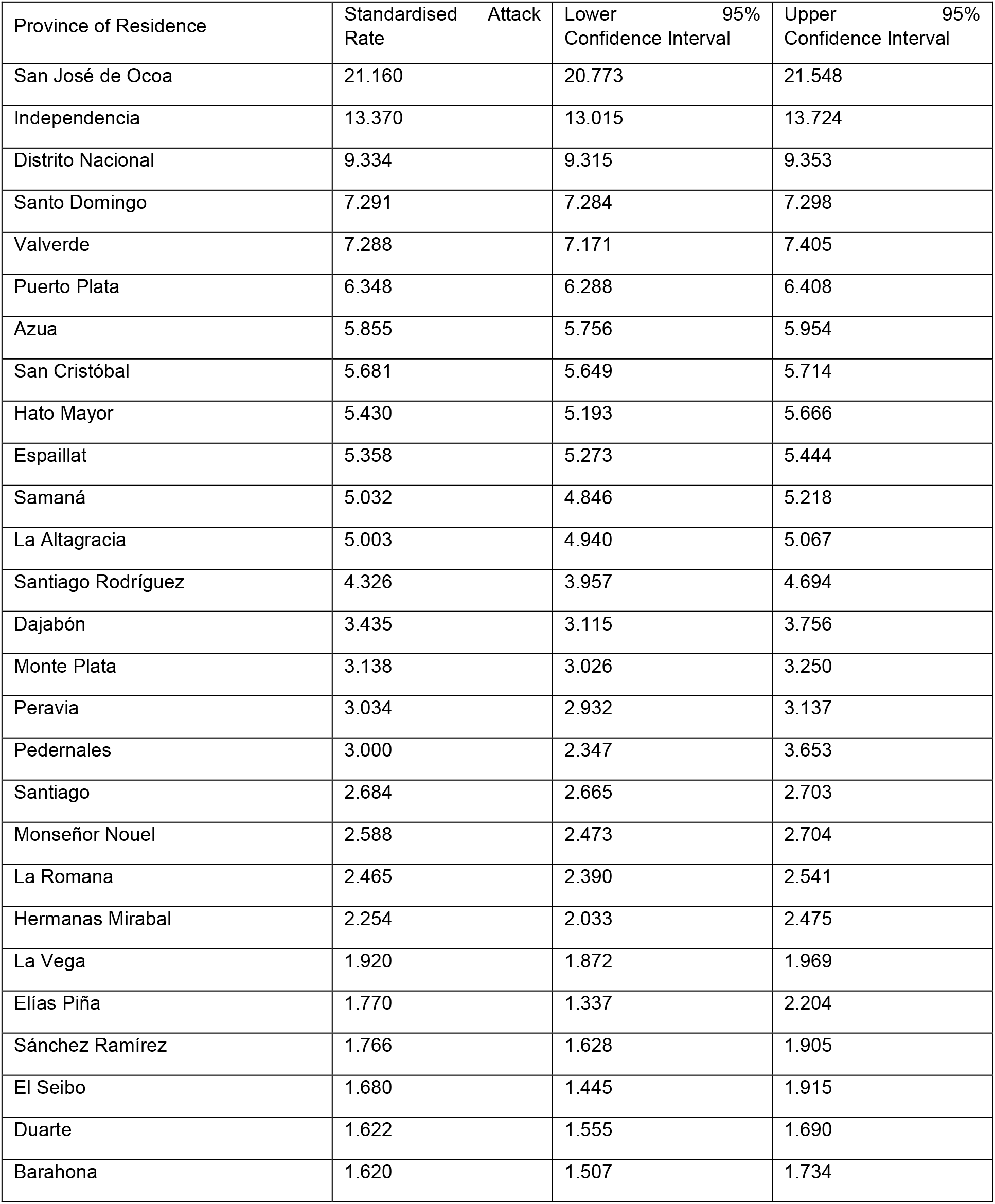
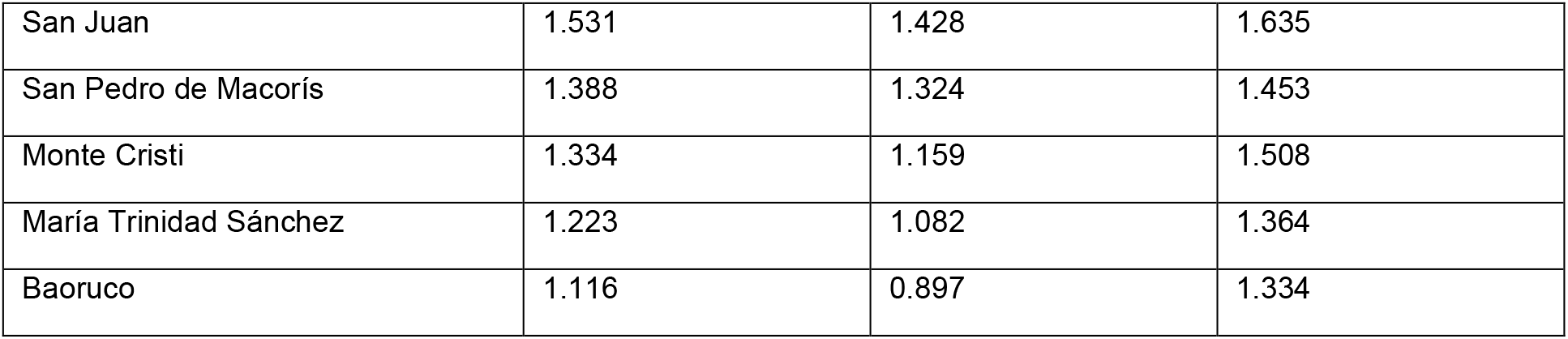
Age and sex standardised attack rates of Zika by province. Respective lower and upper 95% confidence intervals are shown.

**Table S3.**
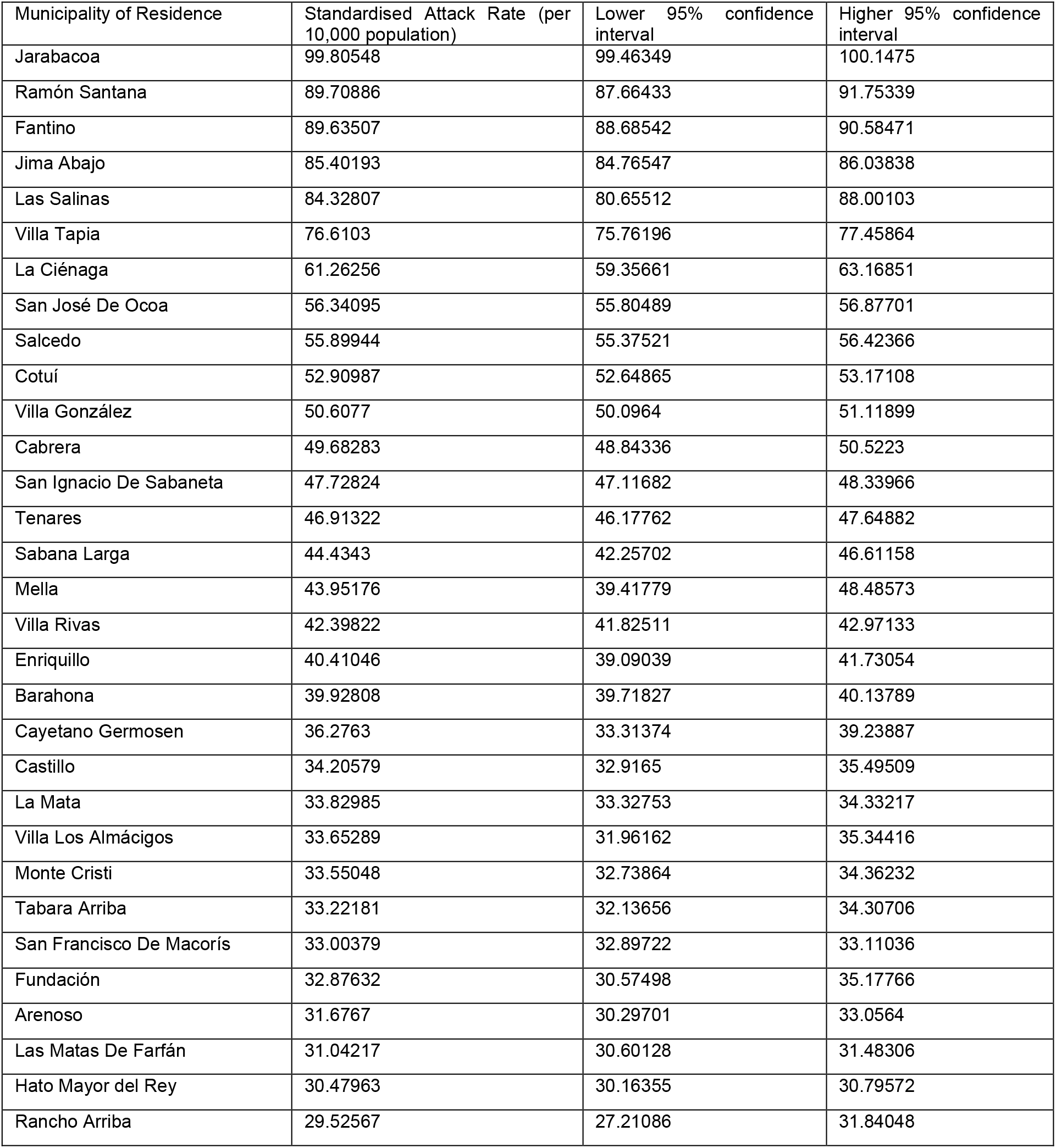
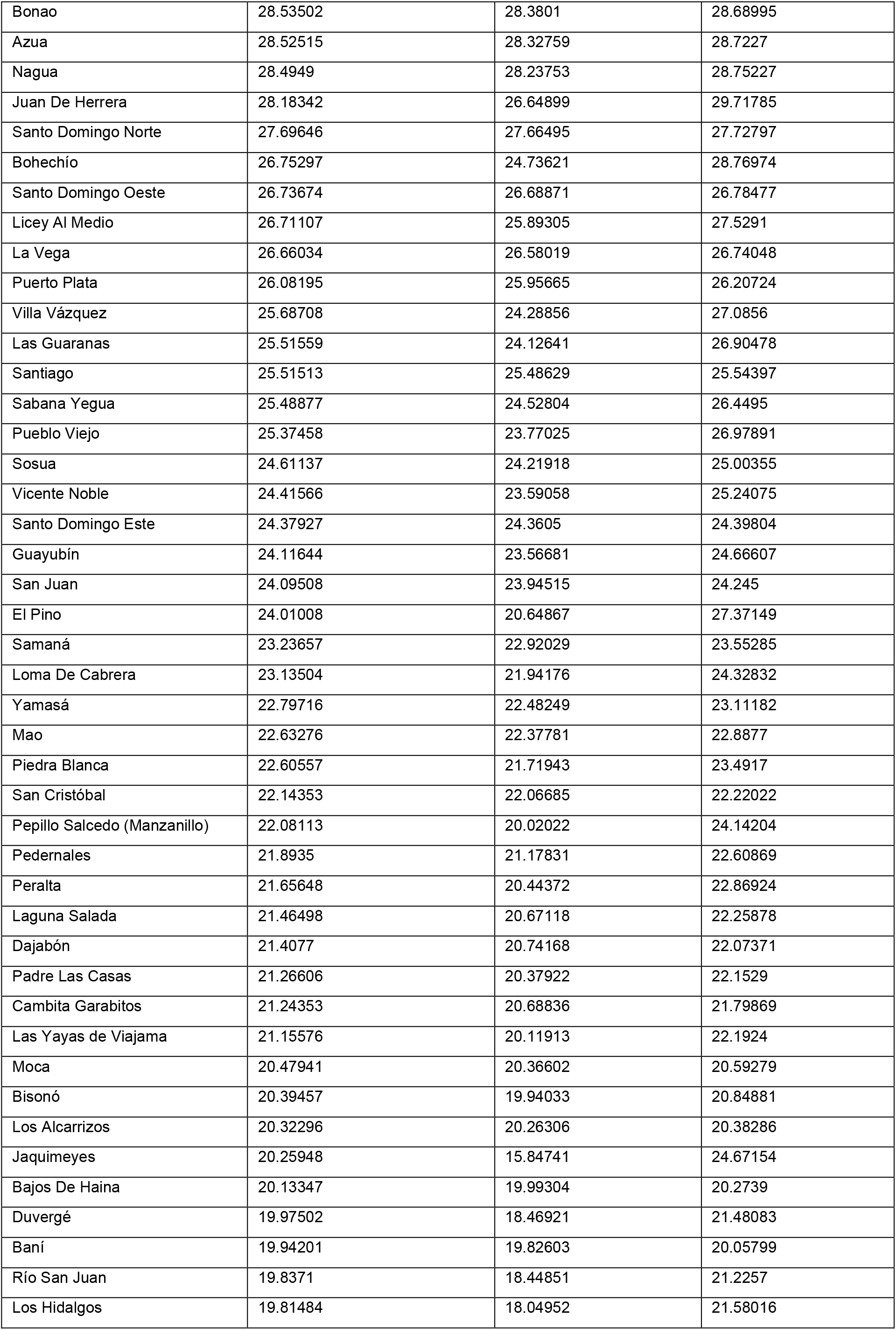
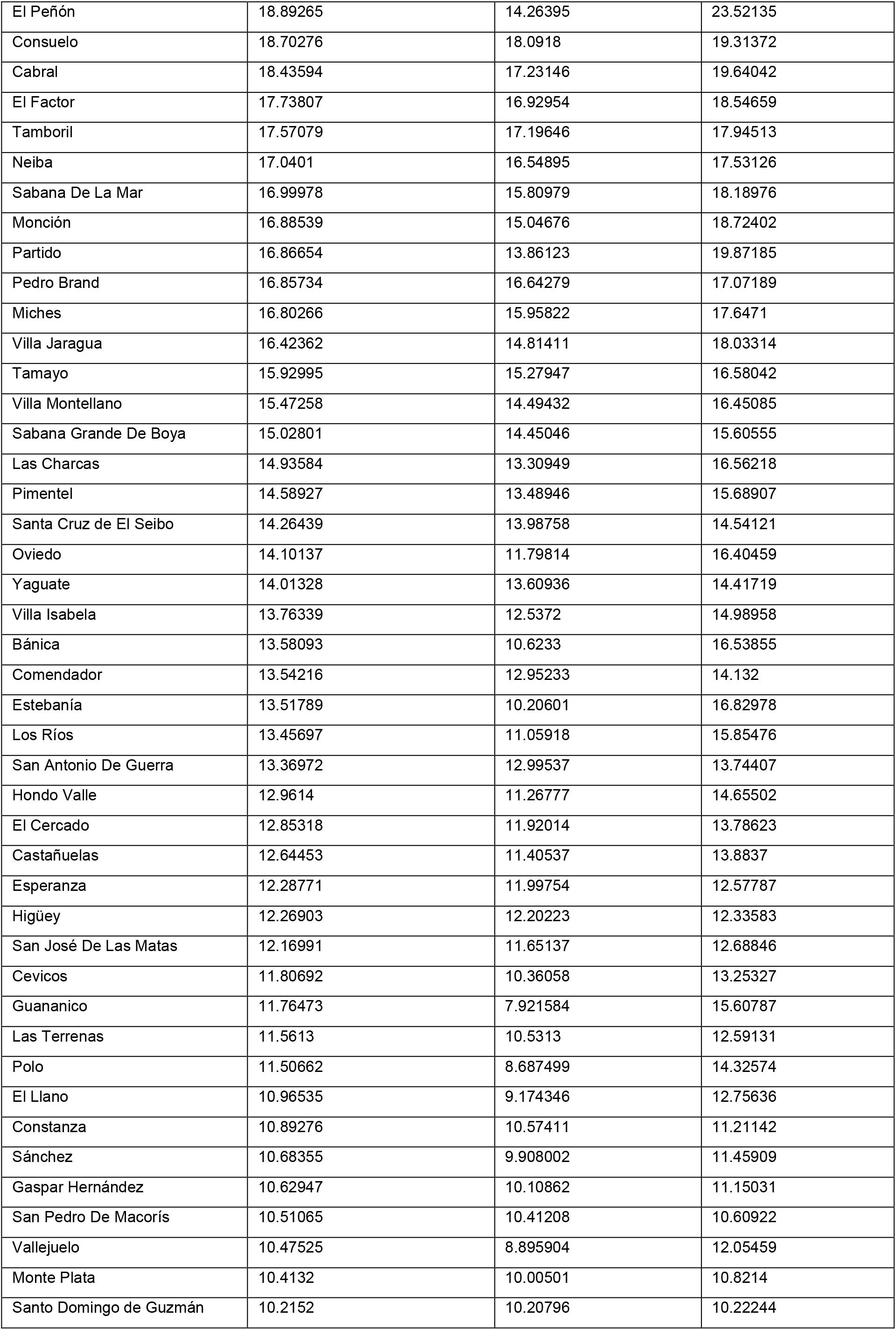
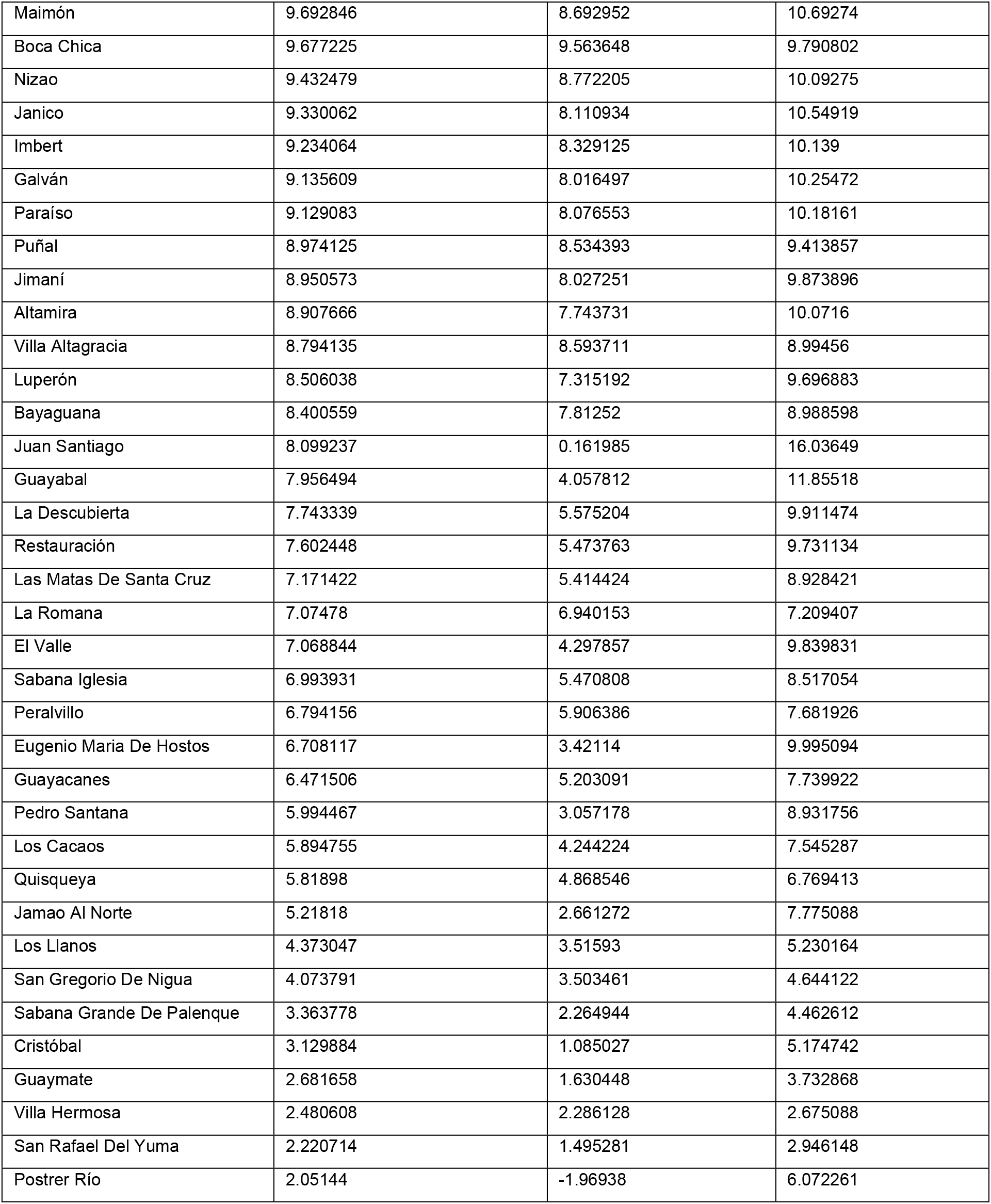
Age and sex standardised attack rates of total dengue by municipality. Respective lower and upper 95% confidence intervals are shown.

**Table S4.**
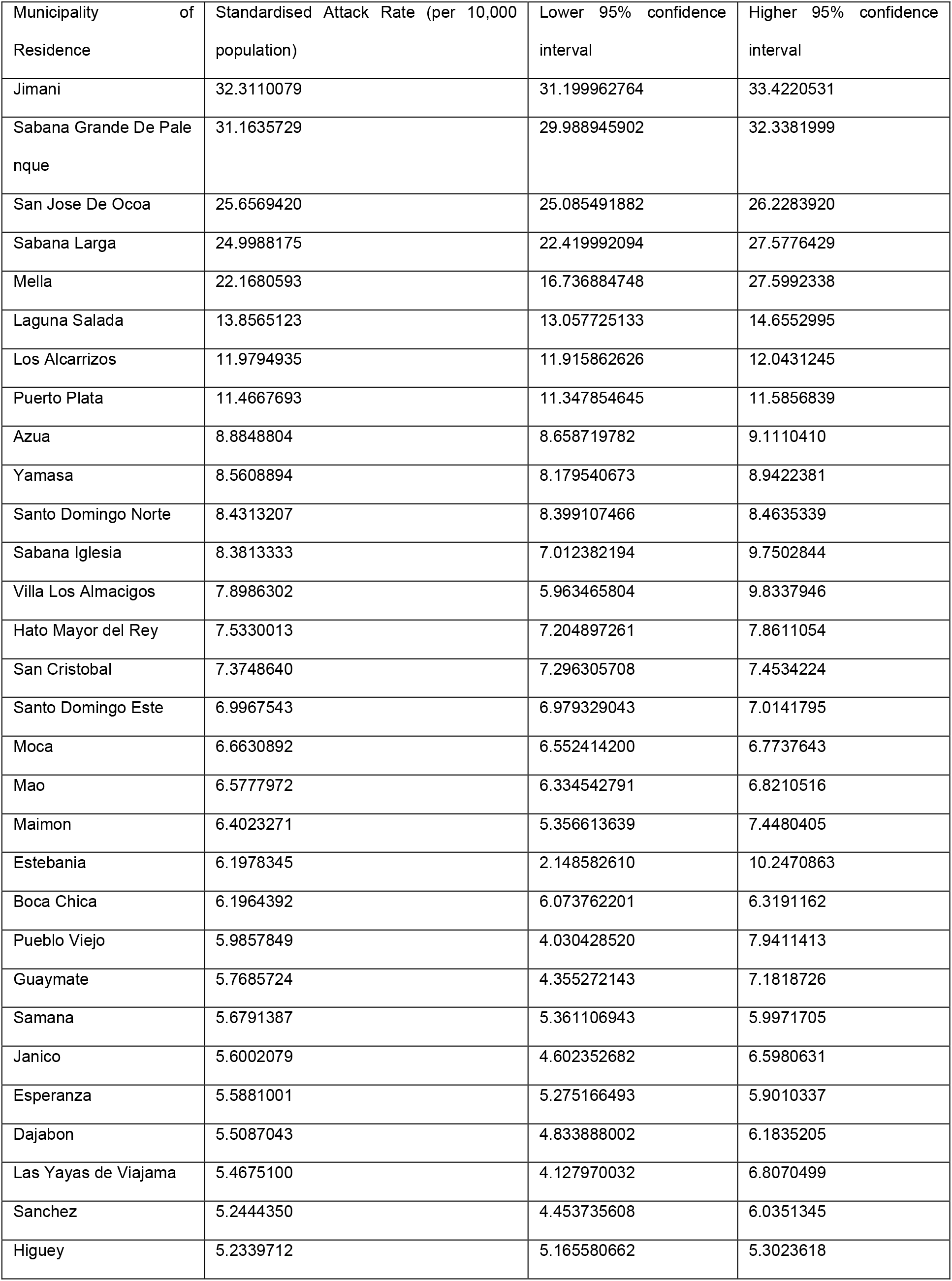
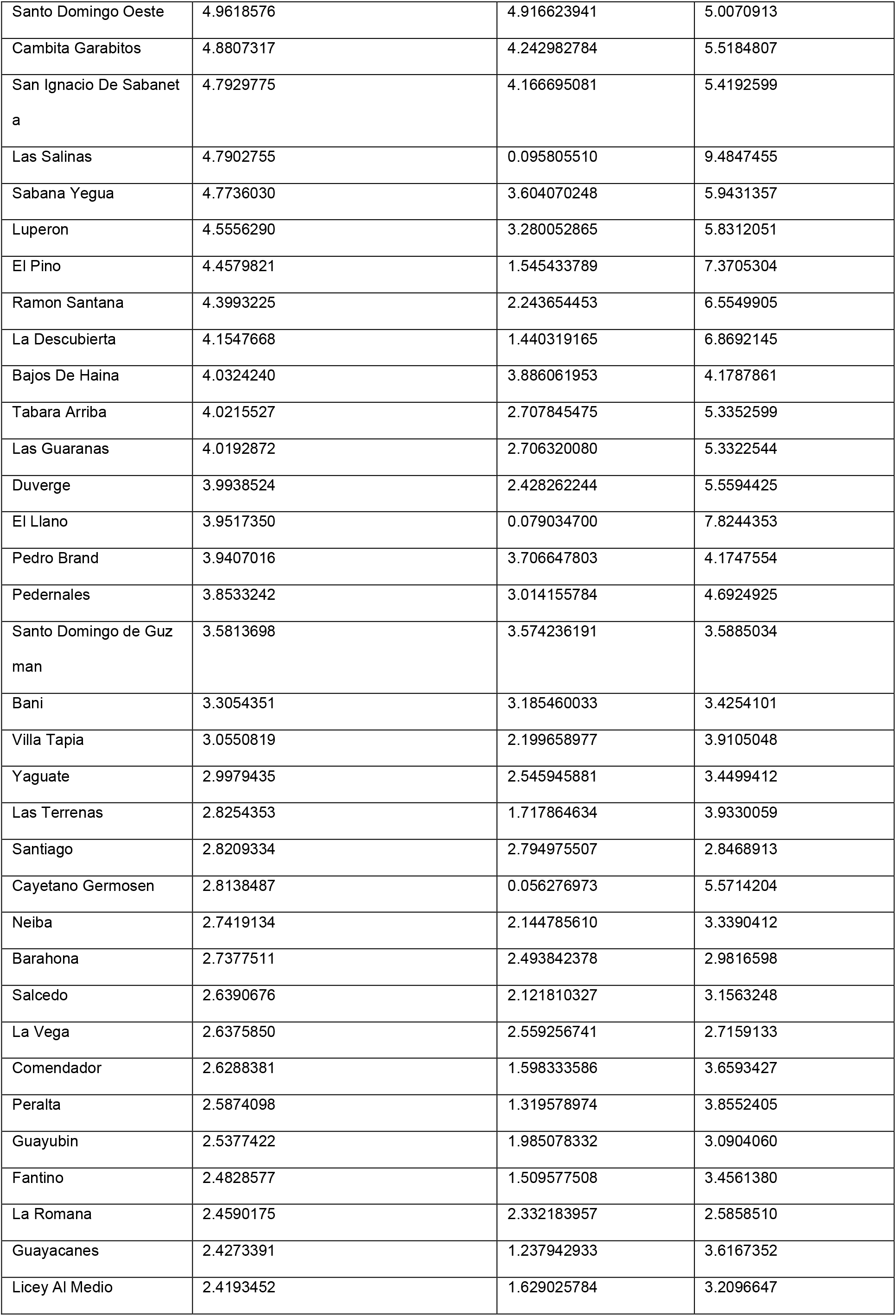
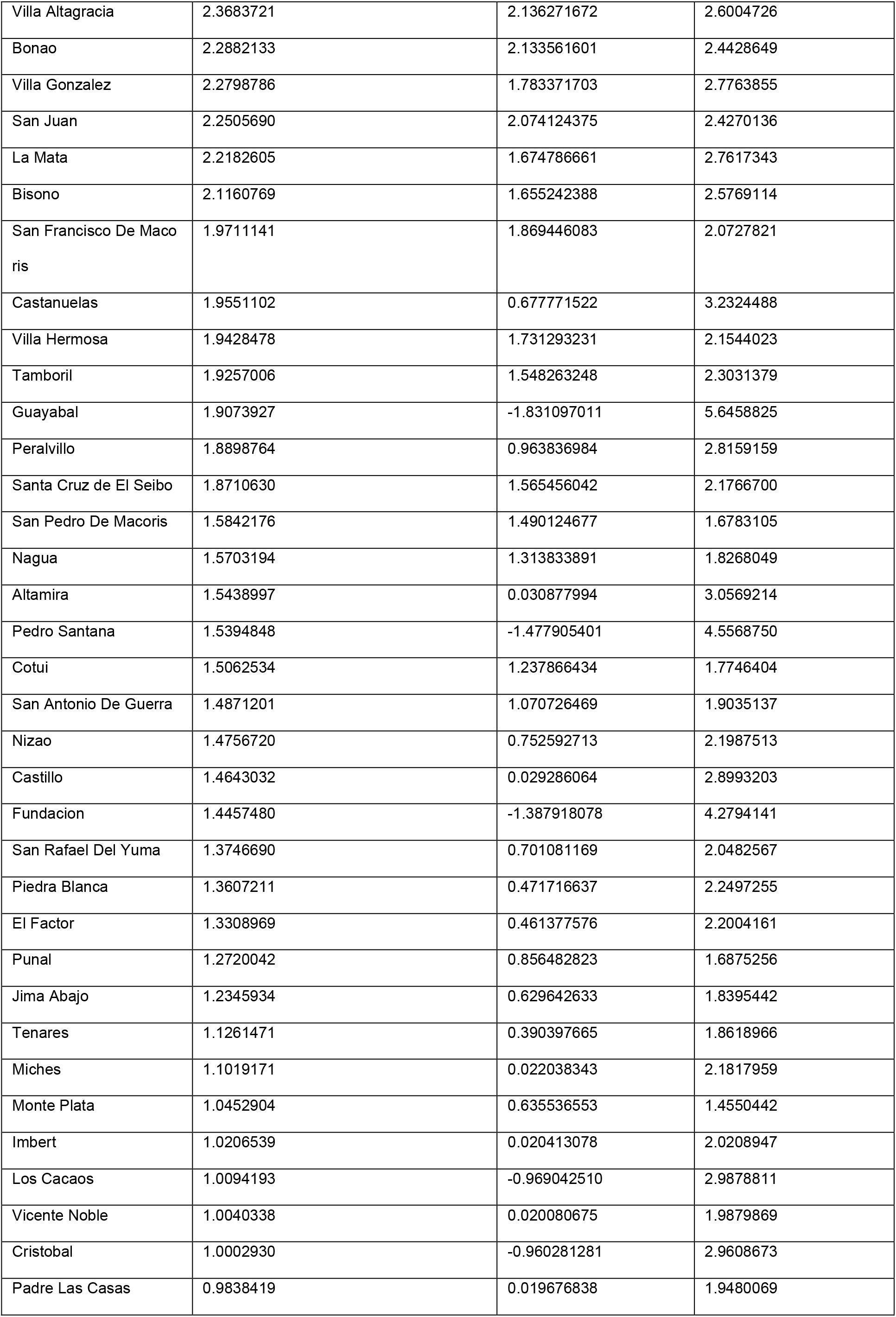
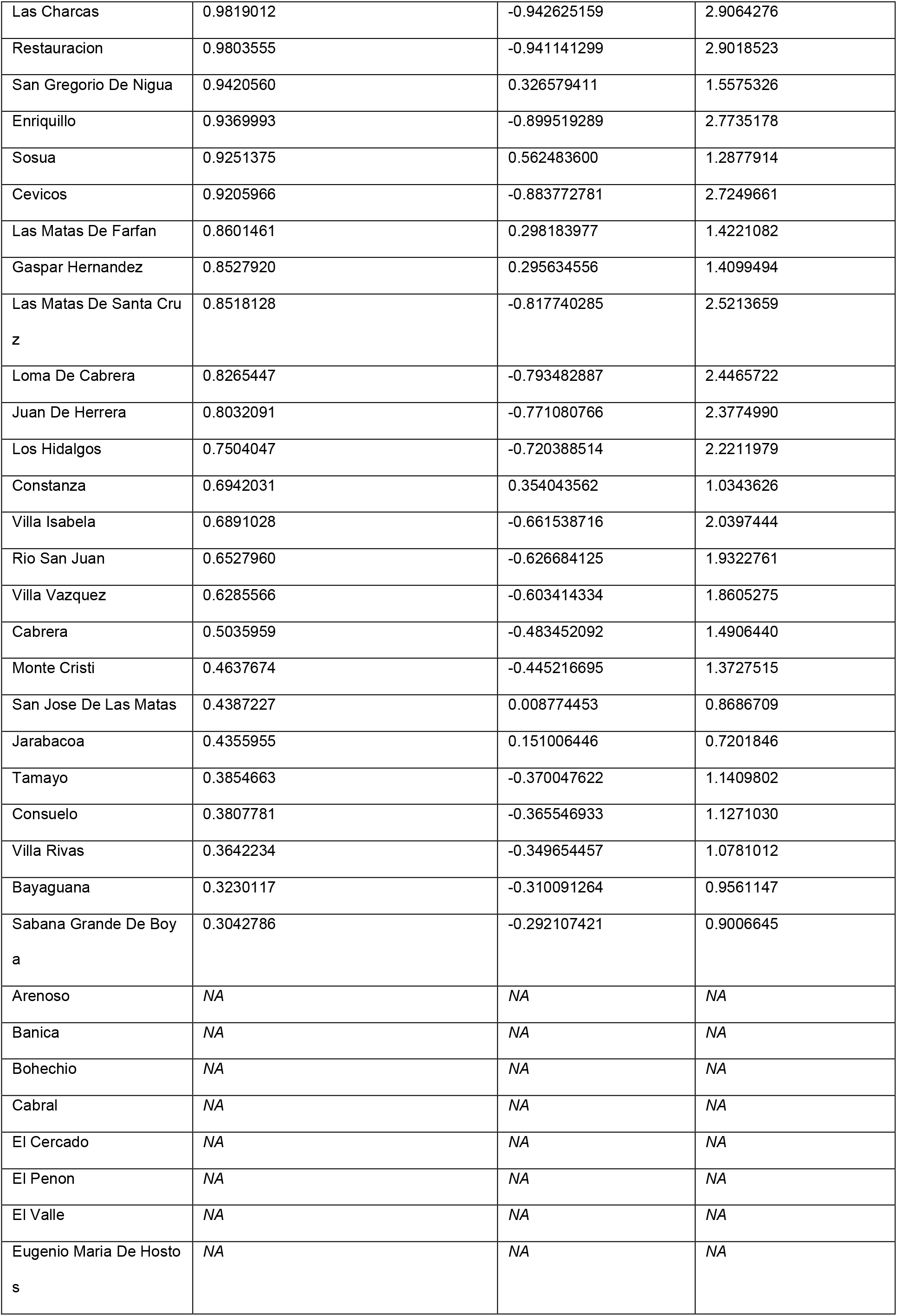
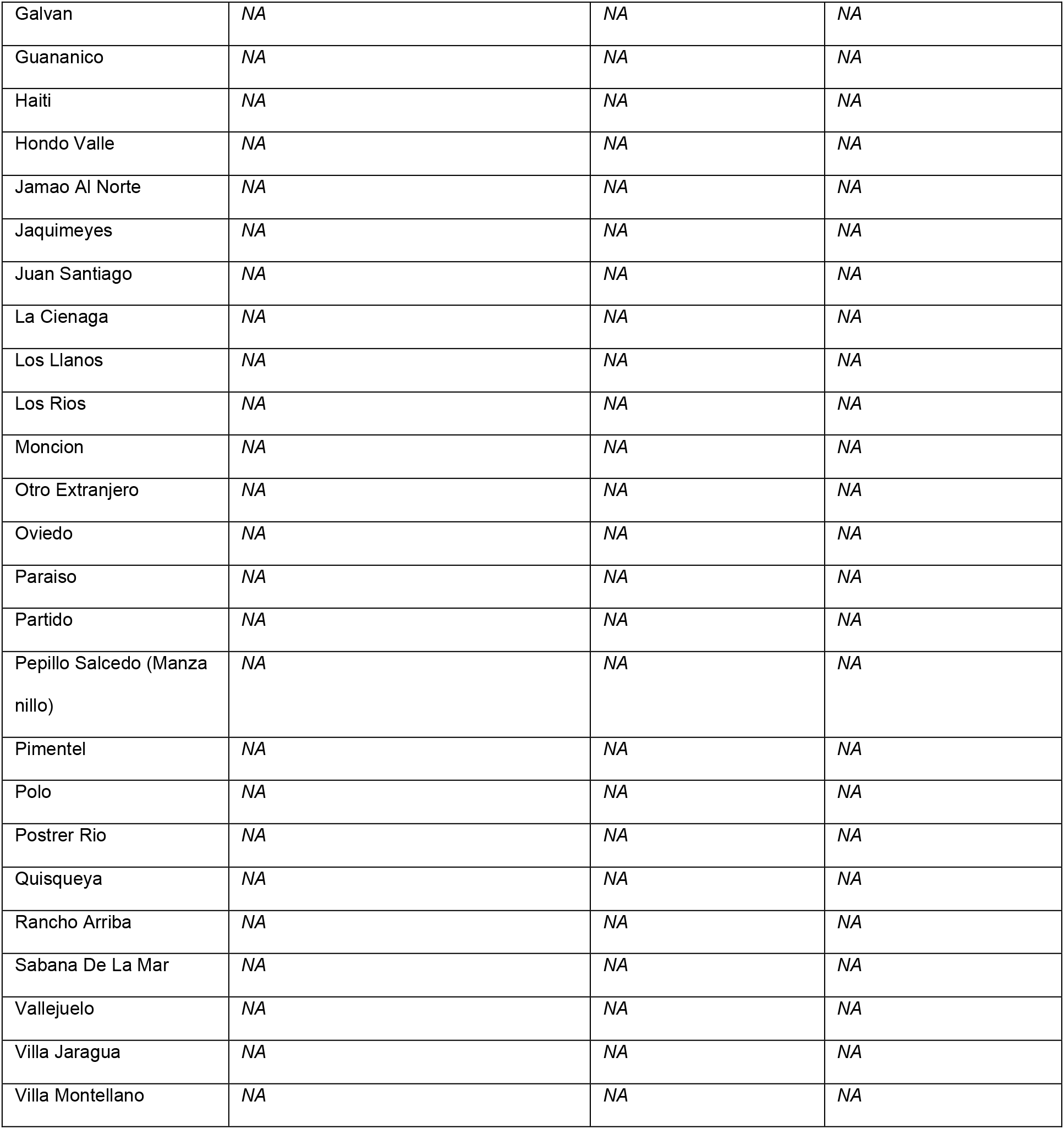
Age and sex standardised attack rates of Zika per municipality. Respective lower and upper 95% confidence intervals are shown.

## References

(1) Davis LE, Beckham JD, Tyler KL. North American Encephalitic Arboviruses. Neurologic Clinics. 2008;26(3):727–57.

(2) Beckham JD, Tyler KL. Arbovirus Infections. CONTINUUM: Lifelong Learning in Neurology. 2015;21:1599–21:1

(3) Bowman LR, Donegan S, McCall PJ. Is Dengue Vector Control Deficient in Effectiveness or Evidence?: Systematic Review and Meta-analysis. PLoS Neglected Tropical Diseases. 2016;10: e0004551.

(4) Bhatt S, Gething PW, Brady OJ, Messina JP, Farlow AW, Moyes CL, et al. The global distribution and burden of dengue. Nature. 2013;496(7446):504–7.

(5) Brady OJ, Gething PW, Bhatt S, Messina JP, Brownstein JS, Hoen AG, et al. Refining the Global Spatial Limits of Dengue Virus Transmission by Evidence-Based Consensus. Reithinger R, editor. PLoS Neglected Tropical Diseases. 2012;6(8):e1760.

(6) Pan American Health Organization, World Health Organization. PLISA Health Information Platform for the Americas. Cases of Zika Virus Disease by Country or Territory. Washington, D.C.: WHO; 2019.

(7) Franz A, Kantor A, Passarelli A, Clem R. Tissue Barriers to Arbovirus Infection in Mosquitoes. Viruses. 2015;7(7):3741–67.

(8) Chye JK, Lim CT, Ng KB, Lim JMH, George R, Lam SK. Vertical Transmission of Dengue. Clinical Infectious Diseases. 1997;25(6):1374–7.

(9) Pomar L, Vouga M, Lambert V, Pomar C, Hcini N, Jolivet A, et al. Maternal-fetal transmission and adverse perinatal outcomes in pregnant women infected with Zika virus: prospective cohort study in French Guiana. BMJ. 2018;k4431.

(10) Counotte MJ, Kim CR, Wang J, Bernstein K, Deal CD, Broutet NJN, et al. Sexual transmission of Zika virus and other flaviviruses: A living systematic review. von Seidlein L, editor. PLOS Medicine. 2018;15(7):e1002611.

(11) Rothman, K., & Greenland, S. (1998). Modern Epidemiology, 2nd Edition. Philadelphia, PA: Lippincott Williams & Wilkins.

(12) admin. Epidemic theory (effective & basic reproduction numbers, epidemic thresholds) & techniques for analysis of infectious disease data (construction & use of epidemic curves, generation numbers, exceptional reporting & identification of significant clusters). Health Knowledge. 2019.

(13) Diekmann O, Heesterbeek JAP, Metz JAJ. On the definition and the computation of the basic reproduction ratio R 0 in models for infectious diseases in heterogeneous populations. Journal of Mathematical Biology. 1990;28(4).

(14) A contribution to the mathematical theory of epidemics | Proceedings of the Royal Society of London. Series A, Containing Papers of a Mathematical and Physical Character. Proceedings of the Royal Society of London. Series A, Containing Papers of a Mathematical and Physical Character. 2019.

(15) Wallinga J. Different Epidemic Curves for Severe Acute Respiratory Syndrome Reveal Similar Impacts of Control Measures. American Journal of Epidemiology. 2004;160(6):509–16.

(16) Last JM, ed. A dictionary of epidemiology. 4th ed. Oxford, England: Oxford University Press, 2001.

(17) Routledge I, Chevéz JER, Cucunubá ZM, Rodriguez MG, Guinovart C, Gustafson KB, et al. Estimating spatiotemporally varying malaria reproduction numbers in a near elimination setting. Nature Communications. 2018;9(1).

(18) Fraser C. Estimating Individual and Household Reproduction Numbers in an Emerging Epidemic. Galvani A, editor. PLoS ONE. 2007;2(8):e758.

(19) Frost WH. Some Conceptions of Epidemics in General. American Journal of Epidemiology. 1976;103(2):141–51.

(20) Abbey H. An examination of the Reed-Frost theory of epidemics. Human biology. 1952;24(3):201–33.

(21) Bowman LR, Rocklöv J, Kroeger A, Olliaro P, Skewes R. A comparison of Zika and dengue outbreaks using national surveillance data in the Dominican Republic. Harley D, editor. PLOS Neglected Tropical Diseases. 2018;12(11):e0006876.

(22) Butler RA. Largest cities in Dominican Republic. Mongabay. Mongabay; 2001.

(23) Dominican Republic administrative boundaries (levels 0-5) - Humanitarian Data Exchange. Humdata.org. 2014.

(24) Wickham. ggplot2: Elegant Graphics for Data Analysis. Springer-Verlag. New York, 2016.

(25) Wikipedia Contributors. Dominican Republic. Wikipedia. Wikimedia Foundation; 2019.

(26) World Bank. Dominican Republic: Country Profile. https://databank.worldbank.org. 2018.

(27) Yamashiro T, Disla M, Petit A, Taveras D, Castro-Bello M, Lora-Orste M, et al. Seroprevalence of IgG specific for dengue virus among adults and children in Santo Domingo, Dominican Republic. The American journal of tropical medicine and hygiene. 2004;71(2):138–43.

(28) WHO (2009) Dengue guidelines for diagnosis, treatment, prevention and control. In: Ciceri K, Tissot P, editors. 2009 ed. Geneva: World Health Organization.

(29) Zika virus disease. World Health Organization. 2016.

(30) Bowman LR, Tejeda GS, Coelho GE, Sulaiman LH, Gill BS, McCall PJ, et al. Alarm Variables for Dengue Outbreaks: A Multi-Centre Study in Asia and Latin America. Hsieh Y-H, editor. PLOS ONE. 2016;11(6):e0157971.

(31) RStudio Team (2018). RStudio: Integrated Development for R. RStudio, Inc., Boston, MA URL http://www.rstudio.com/.

(32) RPubs - Age Adjusted Rates. Rpubs.com. 2017. Available from: https://rpubs.com/bpoulin-CUNY/321735

(33) Justin Lessler, Henrik Salje and John Giles (2018). IDSpatialStats: Estimate Global Clustering in Infectious Disease. R package version 0.3.5. https://CRAN.R-project.org/package=IDSpatialStats

(34) Kempe D, Kleinberg J, Tardos E. Maximizing the Spread of Influence through a Social Network. 2003.

(35) Kilpatrick AM, Randolph SE. Drivers, dynamics, and control of emerging vector-borne zoonotic diseases. The Lancet. [Online] 2012;380(9857): 1946–1955. Available from: doi:10.1016/s0140-6736(12)61151-9 [Accessed: 23rd November 2019]

(36) Energía y Minas en jornada contra dengue junto a líderes comunitarios Cotuí. Energía y Minas en jornada contra dengue junto a líderes comunitarios Cotuí | Presidencia de la República Dominicana. &bsp; 2016.

(37) Kenneson A, Beltrán-Ayala E, Borbor-Cordova MJ, Polhemus ME, Ryan SJ, Endy TP, et al. Social-ecological factors and preventive actions decrease the risk of dengue infection at the household-level: Results from a prospective dengue surveillance study in Machala, Ecuador. Messer WB, editor. PLOS Neglected Tropical Diseases. 2017;11(12):e0006150.

(38) The Relentless Rise of Two Caribbean Lakes Baffles Scientists. Nationalgeographic.com. 2016.

(39) Lednicky J, Beau De Rochars VM, El Badry M, Loeb J, Telisma T, Chavannes S, et al. Zika Virus Outbreak in Haiti in 2014: Molecular and Clinical Data. Reithinger R, editor. PLOS Neglected Tropical Diseases. 2016;10(4):e0004687.

(40) A small town in the Dominican Republic becomes a gateway for relief efforts in Haiti. UNICEF. 2010.

(41) Cosner C, Beier JC, Cantrell RS, Impoinvil D, Kapitanski L, Potts MD, et al. The effects of human movement on the persistence of vector-borne diseases. Journal of Theoretical Biology. [Online] 2009;258(4): 550–560. Available from: doi:10.1016/j.jtbi.2009.02.016 [Accessed: 23rd March 2020]

(42) Lee S, Castillo-Chavez C. The role of residence times in two-patch dengue transmission dynamics and optimal strategies. Journal of Theoretical Biology. [Online] 2015;374: 52–164. Available from: doi:10.1016/j.jtbi.2015.03.005 [Accessed: 23rd March 2020]

(43) 2.3 Dominican Republic Road Network - Logistics Capacity Assessment - Digital Logistics Capacity Assessments. [Online] Logcluster.org. Available from: https://dlca.logcluster.org/display/public/DLCA/2.3+Dominican+Republic+Roa d+Network [Accessed: 23rd April 2020]

(44) Sakkas H, Bozidis P, Giannakopoulos X, Sofikitis N, Papadopoulou C. An Update on Sexual Transmission of Zika Virus. Pathogens. 2018;7(3):66.

(45) Zanluca C, Melo VCA de, Mosimann ALP, Santos GIV dos, Santos CND dos, Luz K. First report of autochthonous transmission of Zika virus in Brazil. Memórias do Instituto Oswaldo Cruz [Internet]. 2015 Jun 9 [cited 2020 Jun 13];110(4):569–72. Available from: https://www.ncbi.nlm.nih.gov/pmc/articles/PMC4501423/

(46) Campos GS, Bandeira AC, Sardi SI. Zika Virus Outbreak, Bahia, Brazil. Emerging Infectious Diseases. 2015;21(10):1885–6.

(47) Ferguson NM, Donnelly CA, Anderson RM. Transmission intensity and impact of control policies on the foot and mouth epidemic in Great Britain. Nature. 2001;413(6855):542–8.

(48) Jombart T, Cori A, Didelot X, Cauchemez S, Fraser C, Ferguson N. Bayesian Reconstruction of Disease Outbreaks by Combining Epidemiologic and Genomic Data. Tanaka MM, editor. PLoS Computational Biology. 2014;10(1):e1003457.

(49) Salje H, Lessler J, Paul KK, Azman AS, Rahman MW, Rahman M, et al. How social structures, space, and behaviors shape the spread of infectious diseases using chikungunya as a case study. Proceedings of the National Academy of Sciences. 2016;113(47):13420–5.

(50) Kucharski AJ, Kwok KO, Wei VWI, Cowling BJ, Read JM, Lessler J, et al. The Contribution of Social Behaviour to the Transmission of Influenza A in a Human Population. Ghedin E (ed.) PLoS Pathogens. [Online] 2014;10(6):e1004206. Available from: doi:10.1371/journal.ppat.1004206 [Accessed: 26th April 2020]

(51) Zika-Epidemiological Report Dominican Republic. 2017.

(52) Zika Virus Outbreak on Yap Island, Federated States of Micronesia | NEJM. New England Journal of Medicine. 2009.

(53) Olliaro P, Fouque F, Kroeger A, Bowman L, Velayudhan R et al. Improved tools and strategies for the prevention and control of arboviral diseases: A research-to-policy forum. PLoS Neglected Tropical Diseases. 2018;12:e0005967.

(54) Bowman LR, Runge-Ranzinger S, McCall PJ. Assessing the Relationship between Vector Indices and Dengue Transmission: A Systematic Review of the Evidence. PLoS Neglected Tropical Diseases. 2014;8: e2848.

(55) Hussain-Alkhateeb L, Kroeger A, Olliaro P, Rocklöv J, Sewe MO, et al. Early warning and response system (EWARS) for dengue outbreaks: Recent advancements towards widespread applications in critical settings. PLoS ONE. 2018;13: e0196811.

